# Artificial Intelligence for Pre-Anaemic Iron Deficiency Detection Using Rich Complete Blood Count Data

**DOI:** 10.1101/2025.06.18.25329494

**Authors:** Daniel Kreuter, Joseph Taylor, Simon Deltadahl, Martin Besser, John R Bradley, Julian Gilbey, Stephen Kaptoge, Nathalie Kingston, Willem H Ouwehand, David Roberts, Olga Shamardina, Kathleen E Stirrups, Dorine Swinkels, Dragana Vuckovic, STRIDES NIHR BioResource, STRIDES Trial, BloodCounts! Consortium, James HF Rudd, Emanuele Di Angelantonio, Carola-Bibiane Schönlieb, Parashkev Nachev, Suthesh Sivapalaratnam, Nicholas S Gleadall, Michael Roberts

**Author notes:** These authors contributed equally to this work (Daniel Kreuter, Joseph Taylor).

## Abstract

Iron deficiency (ID) is a major contributor to global disease burden and the leading cause of anaemia. Early detection is important for proactive management, but conventional complete blood count (CBC) screening often fails to identify non-anaemic iron deficiency (NAID) as many individuals maintain haemoglobin and red cell indices within reference ranges. Analysing 153,565 subjects from the INTERVAL, COMPARE and STRIDES studies, we show that CBC screening has only 40.5 % sensitivity for ID detection, falling to 21.9 % for NAID, highlighting a detection gap. CBC analysers generate a wealth of data beyond Electronic Health Record CBC parameters. We demonstrate that artificial intelligence applied to single-cell flow cytometry data from the CBC analyser achieves 83.7 % sensitivity for ID and 79.3 % for NAID. Our findings show that AI can greatly improve detection of a high-prevalence, important condition without changing infrastructure or diagnostic pathways, providing a valuable tool for proactive anaemia management.

## 1 Introduction

Iron deficiency (ID) is a major contributor to the global burden of disease and the leading cause of anaemia.^1, 2^ Even in the absence of anaemia, non-anaemic iron deficiency (NAID) has emerged as an independent predictor of mortality with increasingly recognised contributions to adverse outcomes across multiple clinical domains, including cardiovascular disease, women’s health, and child development, whilst also serving as an important early indicator in cancer screening.^3, 4, 5, 6, 7^ ID prevalence ranges from 5 % in high socio-demographic index countries to upwards of 35 % in low index countries, and is particularly prevalent in at-risk groups such as women of childbearing age.^8^ The World Bank projects a financial impact of ID amounting to *>* $10 billion each year.^9^ In the UK alone, ID-related hospitalisations have increased by 70 % over five years, contributing to rising healthcare costs of over £30 million annually in secondary care, with primary care costs being substantially higher.^10, 11^ The World Health Organisation (WHO) has recently published a comprehensive framework for anaemia action, including a call for strategies to improve diagnosis.^6, 12, 13^

The ability to detect mild ID anaemia (IDA) and NAID is crucial to early disease identification and efforts to move anaemia management towards a preventative healthcare model, a longstanding focus of health policy.^14, 15^ However, NAID detection is challenging. Conventional screening uses the complete blood count (CBC), but while an individual’s mean corpuscular volume (MCV) and mean corpuscular haemoglobin (MCH) varies little over the course of their lifetime, interpersonal variation is wide.^16, 17^ This leads to large numbers of patients with NAID and early IDA having MCV and MCH values within the CBC reference range, increasing likelihood of missed diagnosis.^18, 19^

We address the diagnostic challenge of early ID and NAID invisibility to conventional screening by leveraging rich data acquired during routine CBC measurement.^18^ During routine CBC acquisition, single-cell level rich CBC (**R-CBC**) data are used to derive standard CBC parameters before being discarded (Fig. 1). We utilise this unprocessed R-CBC data and demonstrate its superior informational content compared to routine Electronic Health Record (**EHR**-)**CBC** parameters for ID detection. To bridge the gap between raw measurements and clinical interpretability, we construct an intermediate data type, High-Dimensional CBC (**HD-CBC**), comprising extended summary reports, analyser messages and quality flags, alongside engineered features derived from the R-CBC measurements. We develop complementary AI approaches optimised for each data type: XGBoost^20^ classifiers for the tabular EHR-CBC and HD-CBC data that enable transparent feature analysis and clinical interpretation, and an end-to-end variational autoencoder (VAE) architecture (**DeepCBC**) that directly processes the complete impedance and flow cytometry signals within R-CBC data. This methodological approach provides independent validation of ID signatures across different data representations while maximising utilisation of the comprehensive measurements routinely captured but typically discarded during standard CBC processing.

**Fig. 1:**
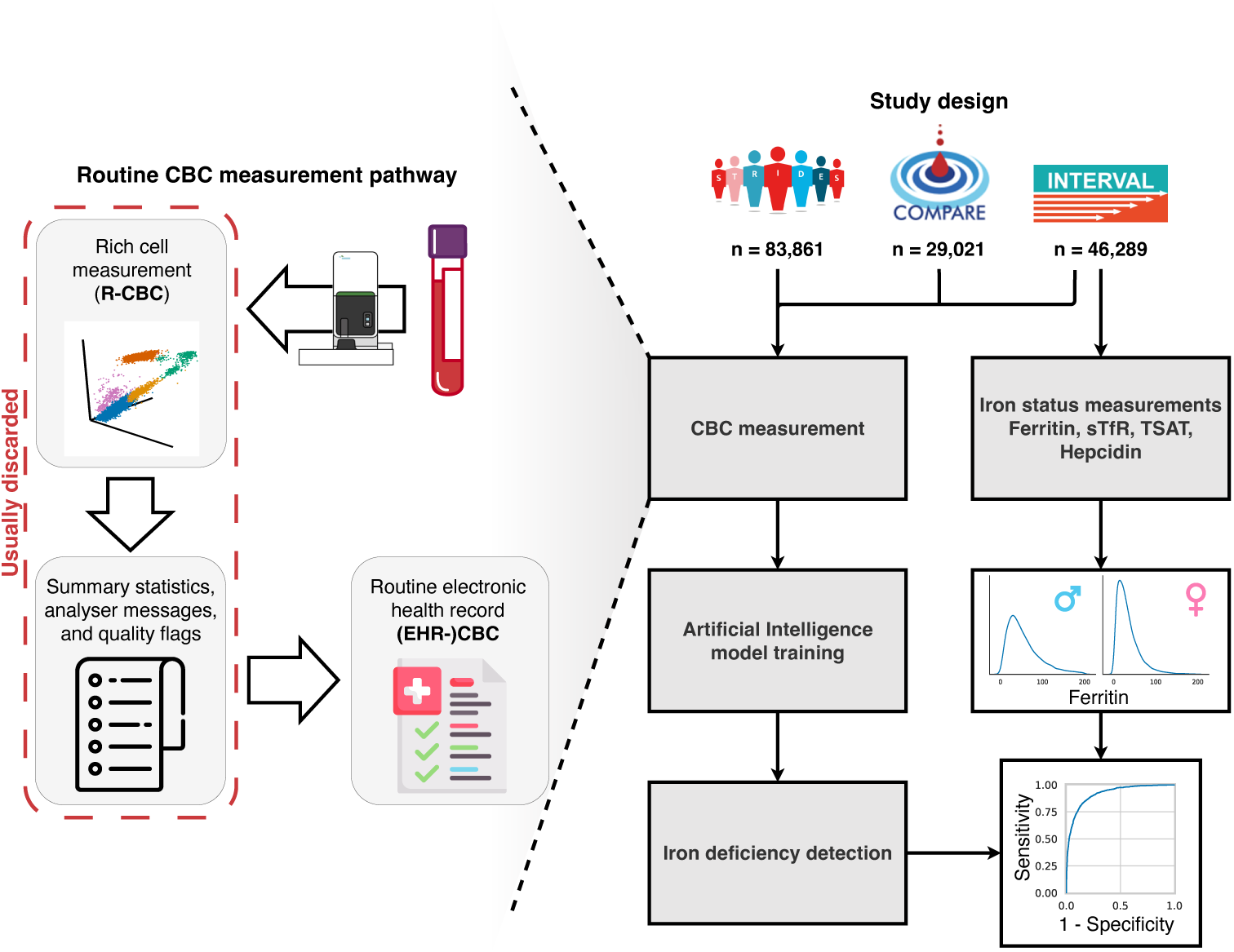
Overview of study design. We leverage the usually discarded rich measurements recorded during routine CBC acquisition (left) from three studies to train AI models to detect ID (right). n, number of individuals; sTfR, soluble transferrin receptor; TSAT, transferrin saturation.

We demonstrate that R-CBC data contains signatures that are known to be difficult for skilled observers to detect from an isolated CBC, including subject sex and blood group. We then apply the models to the task of early detection of ID from an isolated CBC using data from the INTERVAL study, a large randomised controlled trial (RCT) where participants were subjected to regular and measurable iron loss with quantification of iron status, capturing the evolution of NAID. While other works have applied AI approaches to the EHR-CBC, these focused on different tasks, such as distinguishing established IDA from other forms of anaemia or distinguishing ID from haemoglobinopathy traits, or applied rule-based or logistic models.^21, 22, 23, 24, 25, 26, 27, 28^ Our work is, to our knowledge, the first to address the difficult task of detecting ID before the onset of anaemia. Similarly, while elements of HD-CBC and R-CBC data have been utilised in other publications, this is the first time that we are aware of that complete and unaltered R-CBC measurements have been analysed or incorporated into AI models.^29, 30, 31, 32^

We show that the application of AI to expanded CBC data substantially improves the diagnostic power of the CBC test, long regarded as the standard of care for ID screening, and that its use for the detection of NAID far outperforms conventional approaches using CBC reference ranges, without the need for further testing or change to existing pathways for the investigation of anaemia.

## 2 Results

We first demonstrate the additional discriminative power of R-CBC data over the EHR-CBC by the classification of non-disease physiological traits. Secondly, we describe the physiological changes during the evolution of ID as they relate to CBC parameters. Finally, we train and evaluate three models to detect ID from a given CBC and show superior performance with increasing data granularity.

In all analyses, we present results for a held-out test set not used for model training, hyperparameter tuning, or calibration.

### 2.1 Sex and blood group types

To quantify the additional informativeness of R-CBC data, we evaluated the performance of neural network models in classifying biological traits traditionally considered undetectable from standard CBC measurements.^33, 34^ We trained our DeepCBC architecture using R-CBC data from the INTERVAL study and compared its performance against a Multilayer Perceptron (MLP) with identical architecture to DeepCBC’s classification head trained on EHR-CBC data from the same trial. DeepCBC’s VAE component was pre-trained on pooled R-CBC data from the INTERVAL, COMPARE, and STRIDES studies.

DeepCBC consistently outperformed the EHR-CBC model across all classification tasks (Fig. 2b). For sex classification, DeepCBC achieved 93.35 % mean accuracy (95 % confidence interval (CI) = 93.25–93.45 %, t-distribution with 9 degrees of freedom, *n* = 10 independent training runs) compared to 81.94 % (95 % CI = 81.89–81.99 %) for the EHR-CBC model. For blood type classification tasks, the performance differences were more substantial. DeepCBC achieved 59.24 % mean accuracy (95 % CI = 59.11– 59.47 %) compared to near-chance performance of 51.29 % (95 % CI = 51.01–51.57 %) for the EHR-CBC model in differentiating blood group O and non-O blood group (A, B, or AB) individuals. Similarly, for RhD type classification, DeepCBC achieved 63.49 % mean accuracy (95 % CI = 63.22–63.76 %) compared to 50.94 % (95 % CI = 50.71–51.17 %) for the EHR-CBC model. Full confusion matrices including numbers of individuals for all classifications are provided in Extended Data Fig. A1.

**Fig. 2:**
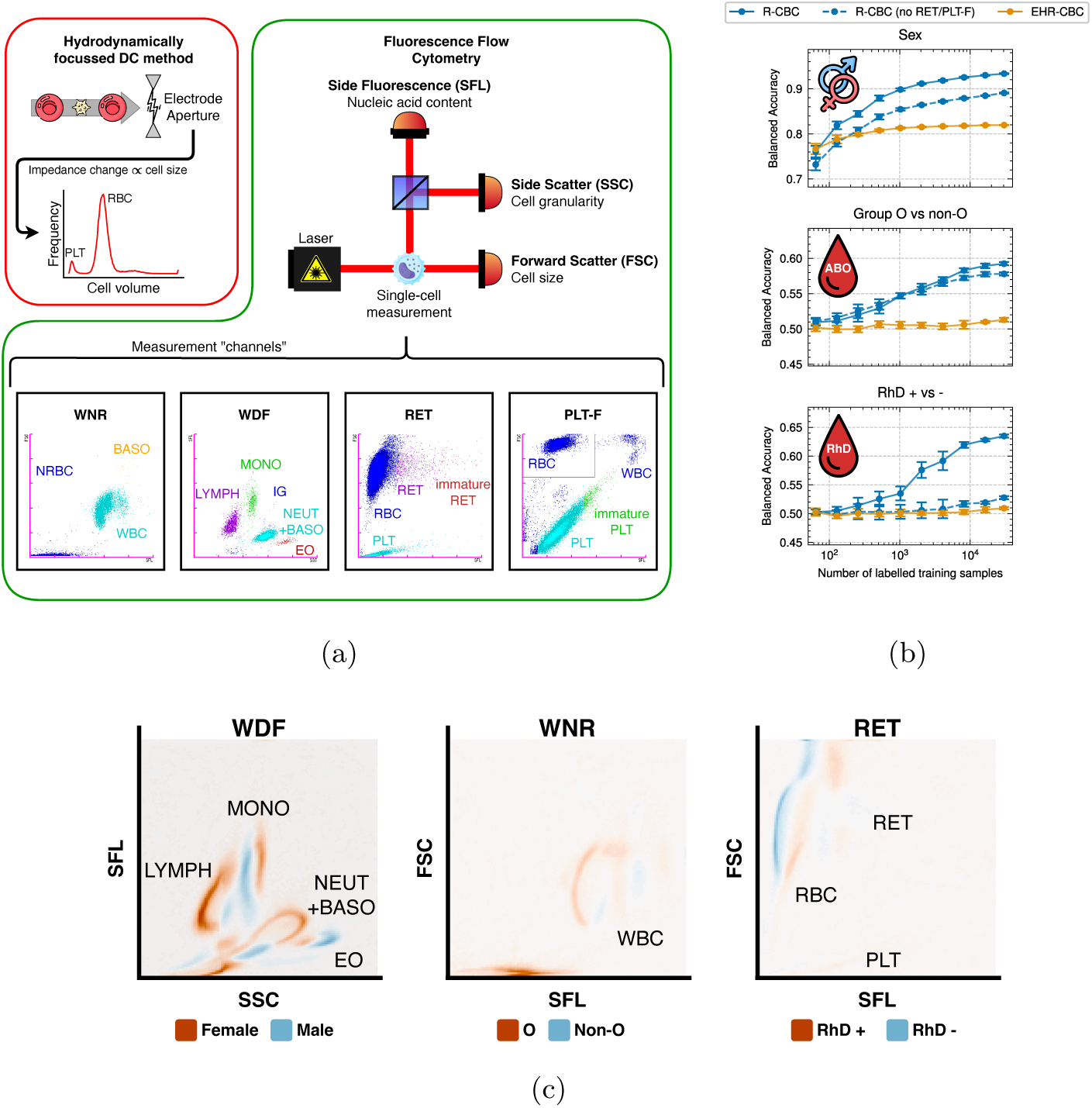
Generation of R-CBC data and evaluation of its informativeness for determination of sex, blood group O vs non-O status, and RhD + vs -. **a,** A sketch illustrating the CBC measurement process inside a Sysmex XN haematology analyser. We call this suite of measurements the R-CBC. The standard EHR-CBC parameters are calculated from the R-CBC. **b,** Performance of neural network models on the classification tasks. Dots and error bars indicate mean and 95 % CI over ten training runs. **c,** Class difference interpolation within R-CBC model. Moving from the negative class (blue) to the positive class (red), density is lost in the blue regions in the scattergram and gained in the red ones. RBC, red blood cells; PLT, platelets; NRBC, nucleated RBC; BASO, basophils; WBC, white blood cells; MONO, monocytes; LYMPH, lymphocytes; IG, immature granulocytes; NEUT, neutrophils; EO, eosinophils; RET, reticulocytes.

To identify which components of the R-CBC data gave rise to these signals, we conducted channel ablation experiments by removing signals from flow cytometry channels used for the enumeration of young red cells (RET channel) and young platelets (PLT-F channel) during training. While removal of these channels did not meaningfully affect classification accuracy for sex or blood group O status (4.29 % and 1.45 % drop), it reduced RhD classification accuracy by 10.72 % to 52.77 % (95 % CI = 52.50–53.04 %), indicating that RET and PLT-F scattergrams contain key information for RhD status detection.

We employed class difference interpolation to visualise changes in the R-CBC data for the three classification tasks (see Methods). Fig. 2c shows the R-CBC channels with the largest differences between classes, exposing marked variation in white cell characteristics between males and females, as well as between group O and non-O subjects. For RhD classification, the side fluorescence signal in the RET channels appears to be the primary discriminating feature learned by the model, consistent with our channel ablation findings.

### 2.2 Evolution of iron deficiency and its relationship to complete blood count parameters

#### Iron status of participants

We analysed 86,340 blood samples from 23,109 male and 23,180 female participants enrolled in the INTERVAL study. Based on the ferritin levels at enrolment, 6,387 (29.2 %) males and 12,436 (56.9 %) females had low iron stores (ferritin between 15– 30 µg*/*L) or ID (ferritin *<* 15 µg*/*L), while 3.5 % of participants were anaemic.^35, 36, 37^ Among participants with 24-month follow-up data, ID prevalence increased substantially in males to 47.3 % (6,472/13,678) and modestly in females to 58.3 % (7,619/13,063).

Over the study period, we observed progressive development of ID and anaemia, with a smaller subset showing recovery from ID (Fig. 3a). NAID was consistently more prevalent than IDA, with NAID:IDA ratios of approximately 7:1 at baseline decreasing to 2:1 at 24 months (Extended Data Table A1).

**Fig. 3:**
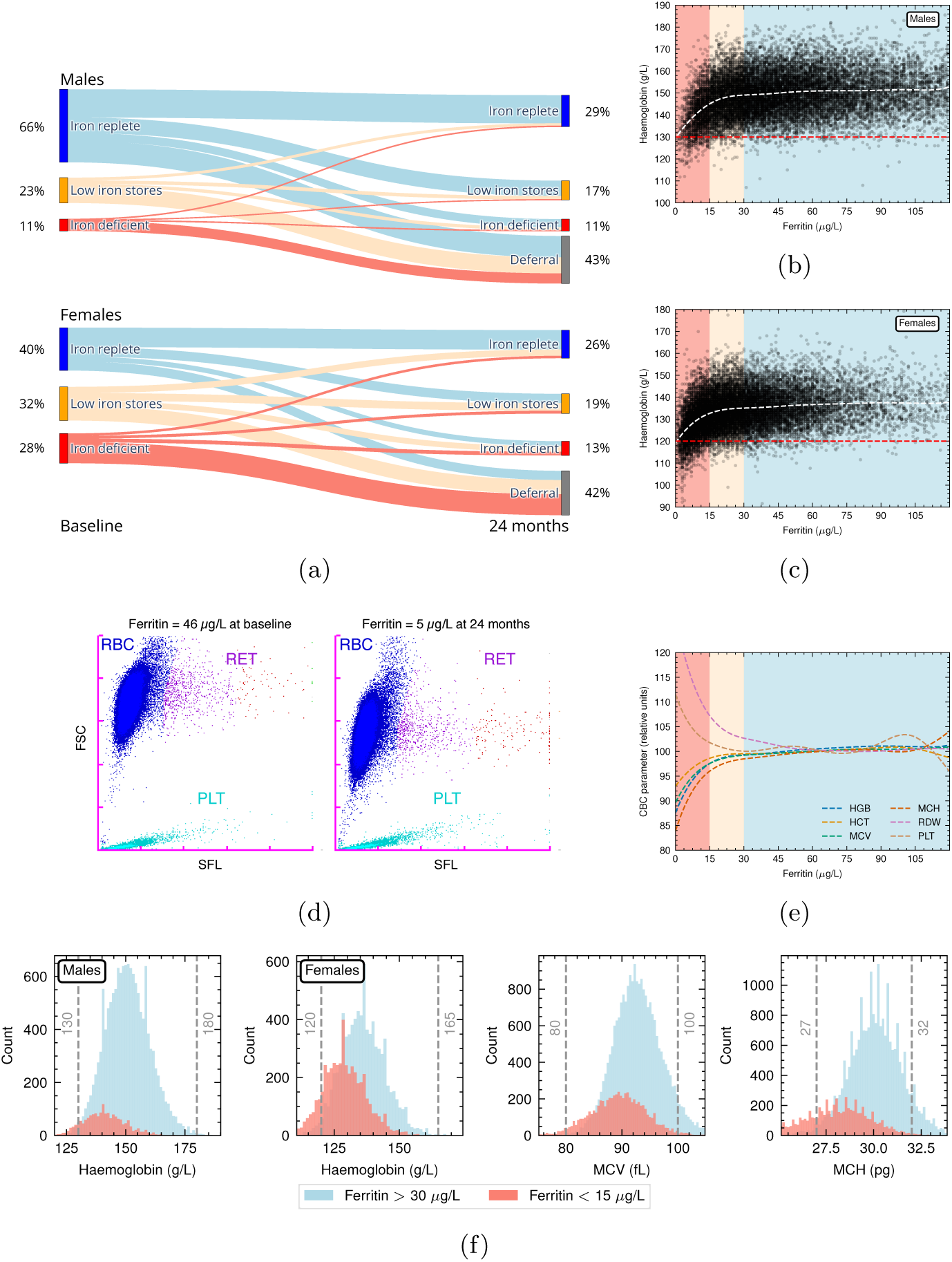
CBC changes with iron depletion. **a,** Sankey plot of changes in subject iron status between baseline and 24 months in INTERVAL for the 8-week male and 12-week female trial arms (shortest donation interval); subjects with at least one low-haemoglobin donation deferral have been separated. **b,c,** Scatter plots of haemoglobin over ferritin for male and female participants at INTERVAL baseline. The background colours indicate iron status as in (**a**). A penalised cubic spline fit of the relationship is indicated as a dashed white line. The red dashed line represents the anaemia threshold. **d,** Example of R-CBC RET channel scattergrams for a blood donor who became iron deficient during the INTERVAL RCT. **e,** Penalised cubic spline fits of multiple CBC parameters against ferritin. Parameters shown in relative units, normalised by their median at ferritin *>* 45 µg*/*L, to enable comparison across different scales. **f,** Distributions of haemoglobin (males and females separated), MCV, and MCH for INTERVAL participants at baseline. Dashed vertical lines indicate CBC reference ranges. WBC, white cell count.

#### Iron deficiency and complete blood count parameters

Analyses of non-linear relationships using penalised cubic splines demonstrated a plateau in the haemoglobin-ferritin relationship in iron-replete participants (ferritin *>* 30 µg*/*L). Iron-replete participants exhibited wide haemoglobin variability (95 % between 133–170 g*/*L in males and 121–153 g*/*L in females) (Fig. 3b, 3c). In the 15–30 µg*/*L range (low iron stores), modest decreases in haemoglobin coincided with lower haematocrit (HCT), MCV, and MCH, and higher red-cell distribution width (RDW) and platelet count (PLT) (Fig. 3e). Below 15 µg*/*L, these parameters showed significant differences compared to participants with ferritin *>* 30 µg*/*L (p *<* 1 *×* 10*^−^*^7^ for all six parameters; independent t-test), demonstrating that CBC parameters respond to ID before the onset of anaemia (see also Extended Data Table A2).

These characteristic changes are illustrated in a male participant who donated 6 units of blood (equivalent to *≈* 1,200 mg of iron^38^) over 24 months during the study (3 donation deferrals). This participant entered with haemoglobin and ferritin levels of 162 g*/*L and 46 µg*/*L respectively, and developed ID with haemoglobin and ferritin of 126 g*/*L and 5 µg*/*L at 24 months (Fig. 3d). Depletion of the subject’s iron stores resulted in a downward shift (reduction of MCV) and lengthening of the red blood cell cluster (increase in RDW).

Despite these characteristic changes, 79.5 % of participants with ID maintained haemoglobin, MCV, and MCH values within reference ranges, highlighting the challenge of ID detection using conventional CBC interpretation (Fig. 3f).

### 2.3 Detecting iron deficiency from complete blood count data using artificial intelligence

In this section we demonstrate the performance of AI classifier models for detecting whether a blood sample belongs to a subject with a ferritin concentration of *<* 15 µg*/*L (indicative of ID, positive class) or *≥* 15 µg*/*L (negative class) using only the subject’s CBC, sex, and age.

#### Comparison of EHR-CBC, HD-CBC, and R-CBC for iron deficiency detection

Performance for ID classification in INTERVAL improved progressively from EHR-CBC to HD-CBC (both using XGBoost), and finally to R-CBC (using our own DeepCBC architecture). The R-CBC model achieved the highest scores across all metrics: F1-score (0.60 in EHR-CBC vs 0.66 in HD-CBC vs 0.66 in R-CBC), balanced accuracy (0.79 vs 0.83 vs 0.83), sensitivity (0.81 vs 0.82 vs 0.84), specificity (0.77 vs 0.83 vs 0.83), positive predictive value (PPV) (0.47 vs 0.54 vs 0.54), receiver operating characteristic area under the curve (ROC AUC) (0.87 vs 0.91 vs 0.91), and average precision (AP) (0.66 vs 0.74 vs 0.75) (Fig. 4c). Using conventional CBC interpretation (ID flagged when any of haemoglobin, MCV, or MCH below reference), sensitivity for ID detection was far below AI model performance at 0.41.

**Fig. 4:**
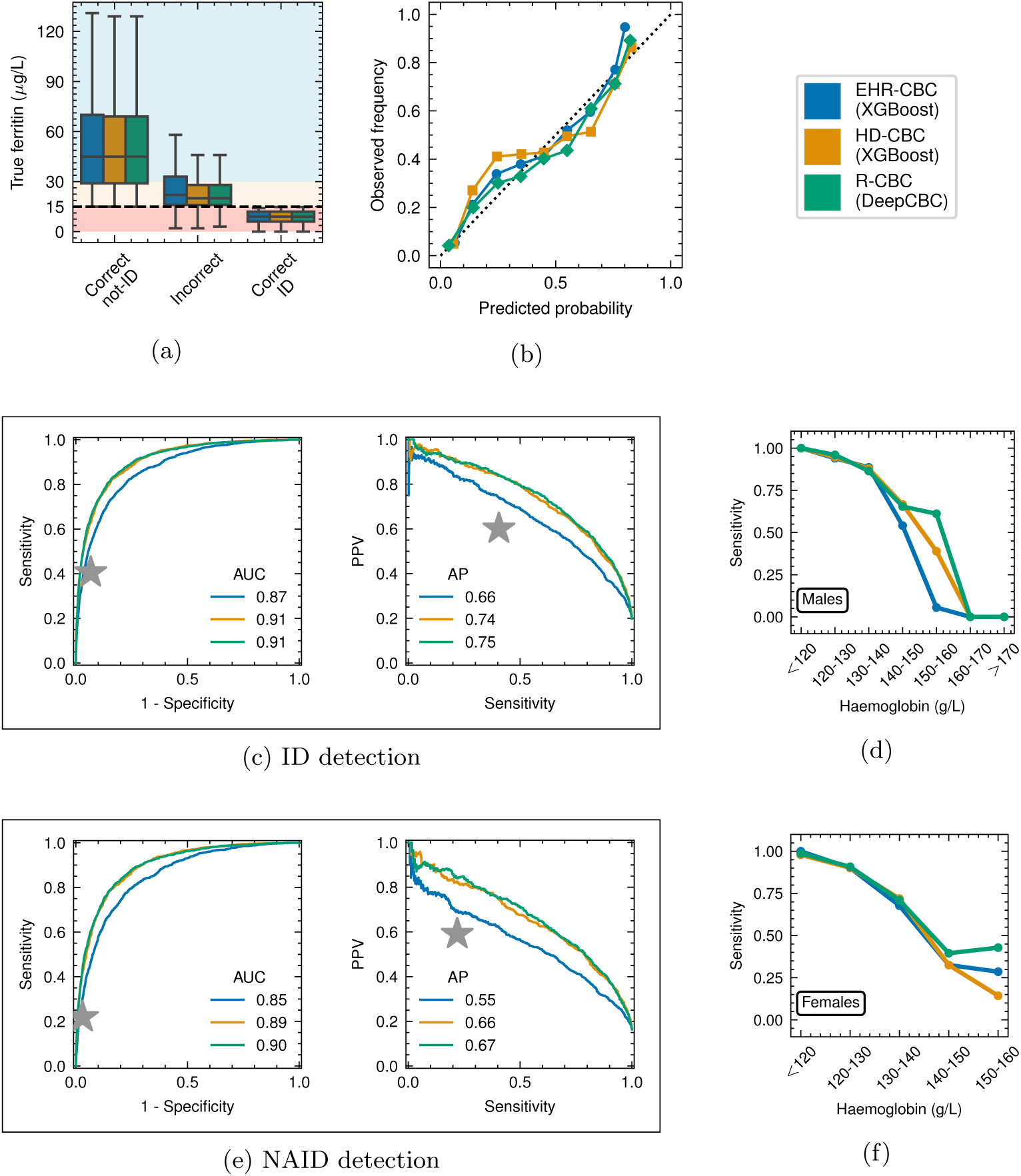
Comparing AI classifier models detecting ferritin. *<* 15 µg*/*L**. a,** Boxplot of true ferritin distribution for the models against the correctness of the models’ prediction. Boxplot whiskers extend to 1.5 IQRs of the data. **b,** Calibration curves (dashed line indicates perfect calibration) for all three models. **c, e,** ROC and PPV-Sensitivity curves, for all samples in the held out test set (**c**) and non-anaemic-only samples (**e**). Stars show performance of conventional reference range approach. **d, f,** Model sensitivity for ID by haemoglobin range, stratified by subject sex. Groups with sensitivity of 0 contained no ID cases. IQR, interquartile range.

When stratified by sex and donation frequency, model performance varied across INTERVAL trial arms (Extended Data Table A3). For male subjects, performance was highest in the 8-week donation interval group (ROC AUC: 0.93 for R-CBC) and slightly lower in the 10-week and 12-week groups. Among female subjects, performance was relatively consistent across donation intervals, with ROC AUCs ranging from 0.886 to 0.893 for R-CBC models.

Models showed similar behaviour in terms of misclassifications and model calibration (Fig. 4a, 4b). Notably, the central 50 % of misclassifications for all three models lie in the 15–32 µg*/*L ferritin range, with HD-CBC and R-CBC models showing slightly narrower ranges.

For our main task of NAID detection, we restricted the test set to non-anaemic samples. In this more challenging task, model performance remained high at AUCs of 0.90 (R-CBC), 0.89 (HD-CBC), and 0.85 (EHR-CBC) (Fig. 4e). Notably, our AI models maintained high sensitivity for NAID detection (79.3 % for R-CBC, 77.6 % for HD-CBC, and 76.2 % for EHR-CBC), outperforming conventional CBC interpretation, which dropped to 22 % sensitivity in the non-anaemic population (Extended Data Table A4).

Evaluating model sensitivity over haemoglobin we found all three models dropping to nearly 50 % sensitivity for ID detection for haemoglobin levels *>* 140 g*/*L, well into the NAID regime (Fig. 4d, 4f). For males, we found that the R-CBC model maintained sensitivity *>* 50 % for haemoglobin levels up to 160 g*/*L.

#### Model explainability

Feature importance analysis revealed distinct patterns between models (Fig. 5a, 5b). The EHR-CBC model primarily relied on conventional red cell indices: MCH, RDW, haemoglobin (HGB), HCT, and MCV. In contrast, the HD-CBC model leveraged advanced parameters absent from the EHR-CBC: the covariance of red cell clusters in the PLT-F scattergram (FSC-SSC diagonal), immature reticulocyte fraction (IRF) in the Y-axis (FSC) of the RET scattergram, haemoglobin, red cell cluster width in the RET scattergram Y-axis (FSC), and percentage of hypochromic red cells (“Hypo-He”). For the image-based DeepCBC model, we illustrate feature importances using class difference interpolation.

**Fig. 5:**
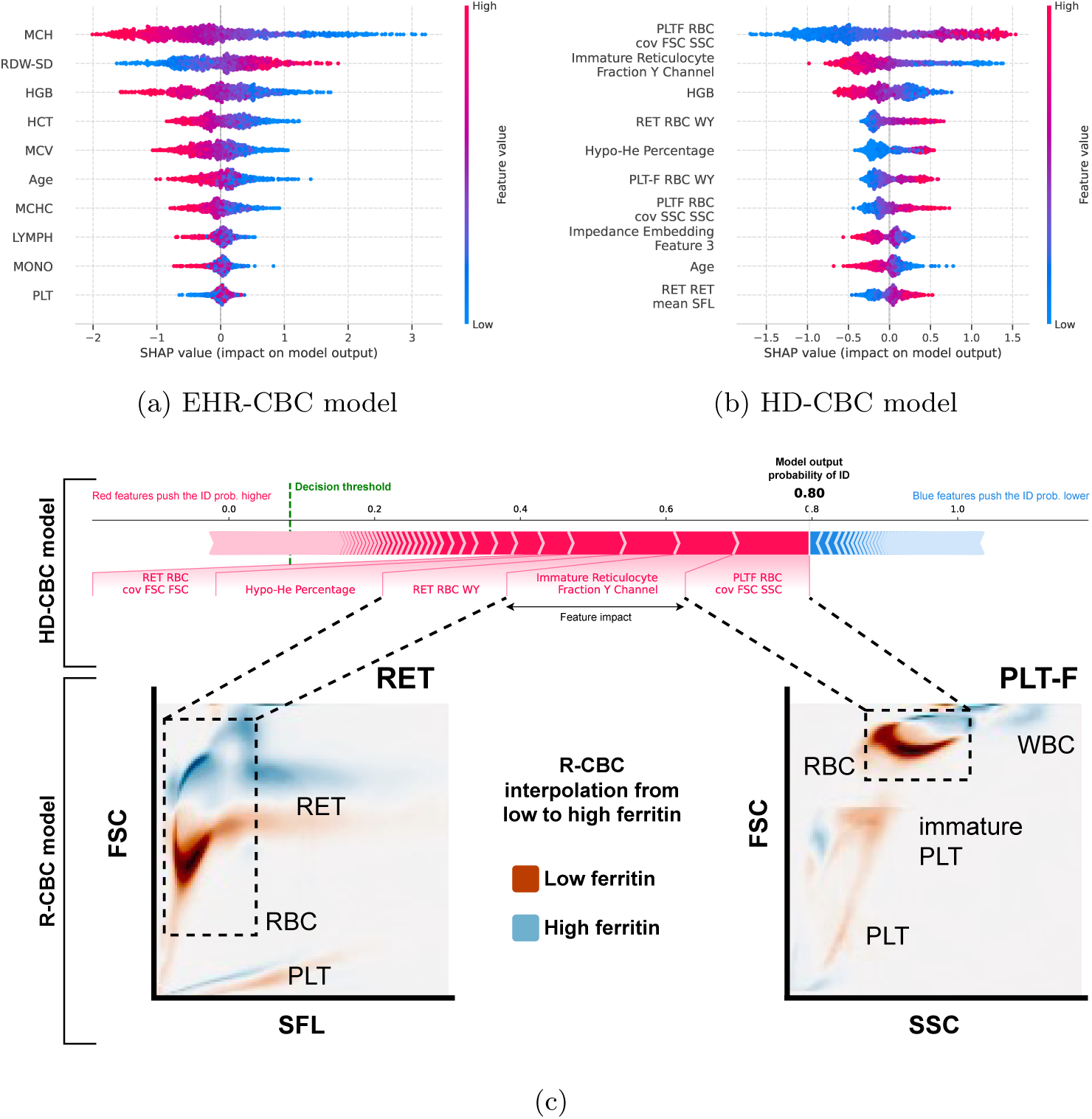
Model explainability for ID detection. **a, b,** SHAP beeswarm plots showing the top ten most important features for the EHR-CBC and HD-CBC models. Positive SHAP value corresponds to increased model output probability of ID. **c,** HD-CBC model single decision explanation for an arbitrary participant in the INTERVAL study with ferritin = 11 µg*/*L (top), and DeepCBC class difference interpolation within the R-CBC RET and PLT-F scattergrams (bottom). For class difference interpolation, moving from low to high ferritin, density is lost in the red regions and gained in blue red ones. Class difference interpolation plots represent the shift in cellular distribution within the cell scattergrams (Fig. 2a). RDW-SD, Red cell distribution width as standard deviation of size distribution; MCHC, Mean corpuscular haemoglobin concentration; LYMPH, lymphocyte count; MONO, monocyte count; cov, covariance; WY, Sysmex-proprietary width in FSC direction.

It is evident that both HD-CBC and R-CBC models independently found crucial differences between ID and iron-replete subjects to be located in the shape of the red cell clusters within the RET and PLT-F R-CBC scattergrams. We illustrate this in Fig. 5c, which shows the explanation of a single decision by the HD-CBC model and class difference interpolation images for the RET and PLT-F R-CBC scattergrams learned by DeepCBC. Both models identify the strongest signal for ID detection as a shift in the red cell cluster width in the FSC-SSC diagonal direction; i.e., a reduction in the size and complexity or content of the red cells as measured within these channels.

#### Investigating model predictions

To gauge our approach’s limitations, we investigated misclassified samples for our best-performing model, the R-CBC-based DeepCBC architecture.

First, we examined model and conventional CBC screening predictions over haemoglobin and ferritin. Model performance markedly exceeded conventional CBC screening for NAID (Fig. 6a, 6b). Again, we observe false positives by the model clustering close to the ferritin = 15 µg*/*L threshold.

**Fig. 6:**
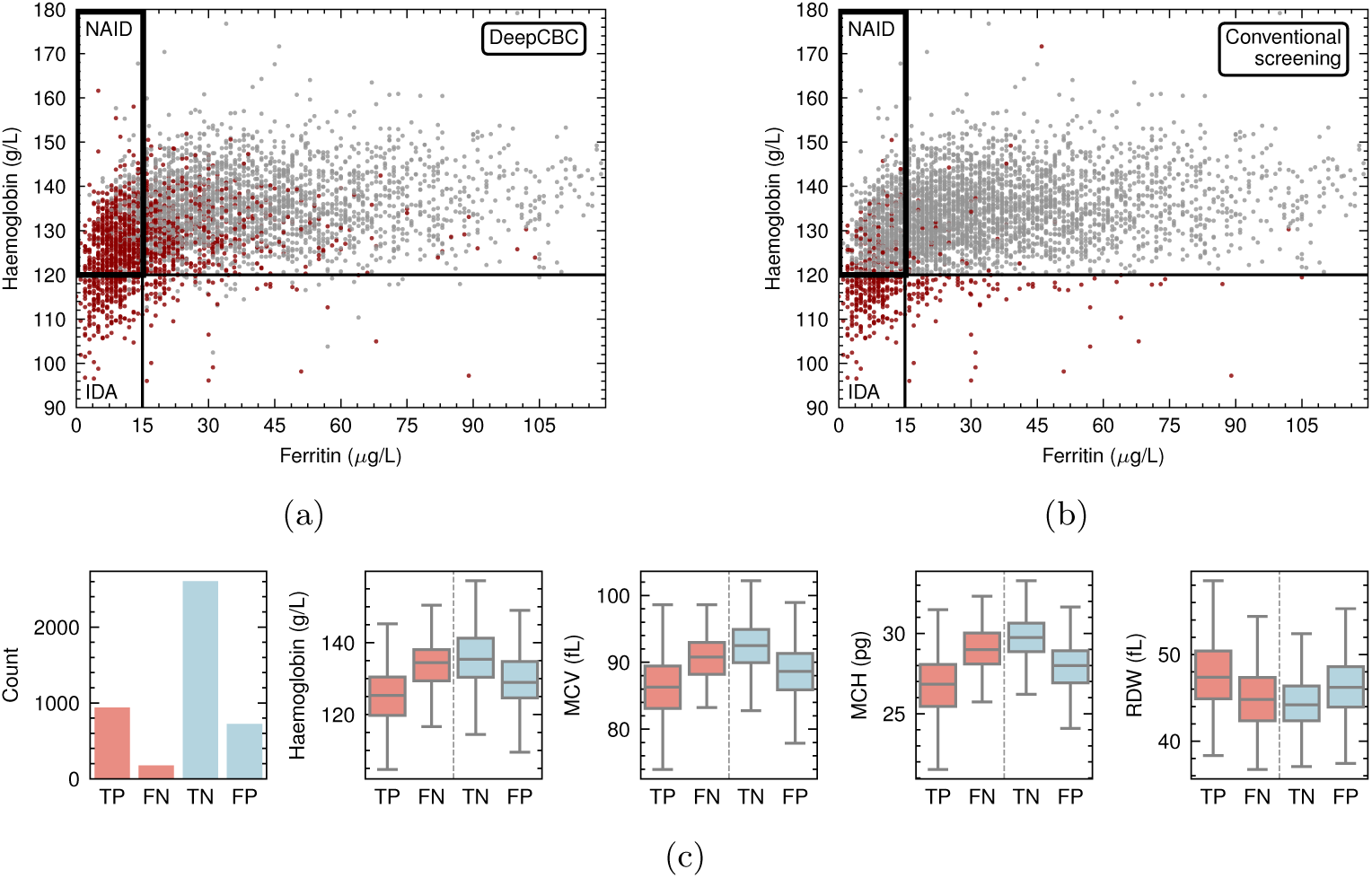
Investigation of R-CBC model predictions. **a, b,** Haemoglobin versus ferritin scatter plots for female subjects in the held-out test set, with iron deficiency predictions shown in red and non-iron deficiency predictions in grey. DeepCBC predictions (**a**) and conventional CBC screening predictions (**b**). **c,** Distributions of haemoglobin, MCV, MCH, and RDW by model classification outcome. Boxes indicate IQR with whiskers extending to outermost data points within 1.5 IQR from box. TP, true positives; FN, false negatives; TN, true negatives; FP, false positives.

Looking further at DeepCBC predictions, we observed key CBC parameters differing significantly between misclassified and correctly classified samples, generally showing lower haemoglobin, lower MCV, lower MCH, and higher RDW in true positives versus false negatives: median haemoglobin was 128.0 g*/*L in true positives vs 136.4 g*/*L in false negatives; median MCV was 85.2 fL vs 89.7 fL; median MCH was 26.5 pg vs 28.8 pg; and median RDW was 46.4 fL vs 44.9 fL (p *<* 0.0001 for all comparisons, Mann–Whitney *U* test; Fig. 6c).

Finally, recognising that while ferritin concentration is the most widely used biomarker of iron status it is not a perfect marker of iron stores, we sought to validate model performance against other clinically validated iron biomarkers. Evaluation of model predictions against transferrin saturation, soluble transferrin receptor (sTfR), sTfR to log-ferritin ratio, and hepcidin showed analogous behaviour to ferritin, particularly in the case of sTfR to log-ferritin ratio (Extended Data Fig A5).

## 3 Discussion

This study demonstrates that the application of artificial intelligence (AI), augmented by the use of rich complete blood count (R-CBC) data, can substantially improve on current standard of care for iron deficiency (ID) screening. Moreover, this is achieved without the need for changes to existing infrastructure or care pathways, and reduces the need for further laboratory testing. ID is a major public health challenge in both lowand high-income countries, and a leading contributor to the global burden of disease.^8, 1^ We show that screening using standard CBC reference ranges fails to identify 79.5 % of non-anaemic ID (NAID) cases, delaying diagnosis. Our study demonstrates that AI models applied to rich data that is routinely recorded but typically discarded during CBC acquisition can improve screening sensitivity for non-anaemic ID (NAID) from 21.9 % to 79.3 %.

The current study differs fundamentally from previous studies applying AI methods in the field of ID and anaemia in three aspects: task, method, and scale. Firstly, while previous studies tackle the task of separation of established ID anaemia from other forms, or the separation of ID from thalassaemia traits,^24, 28, 25, 26, 27^ our focus is instead on the identification of early, pre-anaemic ID with the goal of preventative management. Secondly, we differ in method, modelling rich CBC data. This is the first study, to our knowledge, to apply AI to this data type. Rich CBC data enabled us to develop novel, highly accurate models. We have previously demonstrated that such data contains valuable information for predicting genetic factors influencing platelet reactivity and blood cell traits.^29, 31^ Finally, we significantly increase the scale of datasets used for training and validation of ID AI models. The largest known study, by Kurstjens et al.,^24^ used EHR-CBC data from 11,818 subjects. In the INTERVAL dataset alone we have data for 46,289 participants and we pre-train DeepCBC using data from 153,565 participants from the INTERVAL, COMPARE, and STRIDES studies. This scale and the comprehensive iron testing within INTERVAL provided an unprecedented opportunity to develop robust AI models for early ID detection.

To quantify the informational gain of rich CBC data, we first demonstrated its capacity to accurately classify biological traits traditionally considered undetectable from standard blood count measurements. Our DeepCBC model achieved 93.4 % accuracy in sex classification, substantially outperforming a model with similar classifier architecture using only the standard CBC parameters (81.9 % accuracy). Furthermore, the model could detect subtle differences in blood cell characteristics associated with blood group O status (59.2 % accuracy) and RhD type (63.5 % accuracy) without any direct measurements of these antigens. When we conducted channel ablation experiments by removing reticulocyte (RET) and optical platelet (PLT-F) R-CBC flow cytometry channels, classification accuracy for RhD type dropped to a random chance level, while sex and blood group O classification remained largely unaffected. This finding aligns with biological understanding, as RhD proteins are exclusively expressed on red cell membranes with RhD mRNA upregulation occurring during the reticulocyte stage, while ABO antigens are present across all blood cell lineages.^39^

Our dataset contained a high proportion of non-anaemic ID cases (14.5 % at baseline, increasing to 15.7 % at 24 months), providing excellent training data for this otherwise challenging diagnostic scenario and avoiding bias toward established ID anaemia typically found in clinical datasets. In line with previous studies, we found large variation in haemoglobin levels in iron-replete participants (ferritin *>* 30 µg*/*L), representing inter-personal variability in physiological baseline.^17^ Penalised cubic spline analysis also showed that the large numbers of participants with NAID have haemoglobin levels below their physiological normal level despite not reaching thresholds for anaemia, demonstrating that NAID has measurable physiological impact even before anaemia development. The clinical significance of this is supported by well-documented independent associations between NAID and mortality.^4, 5^

For the primary task of ID detection, both our high-dimensional CBC (HD-CBC) XGBoost and rich CBC (R-CBC) DeepCBC models demonstrated excellent performance, with balanced sensitivities between sexes. Compared to conventional blood count interpretation using haemoglobin, mean corpuscular volume (MCV), and mean corpuscular haemoglobin (MCH) standard reference ranges, our models achieved substantially higher sensitivity while maintaining specificity. For non-anaemic ID detection in particular, the DeepCBC model maintained 79.3 % sensitivity compared to 21.9 % for conventional interpretation.

Our dual modelling approach allowed for independent validation of biological feature importance. While models based on the conventional CBC primarily relied on established red cell indices (MCH, red cell distribution width, and haemoglobin), our enhanced models leveraged more sophisticated parameters. The HD-CBC and R-CBC models independently found red cell cluster shapes in flow cytometry scattergrams in the forward scatter (cell size) and side scatter (cell content) directions to be the most important factors in ID detection, demonstrating that the measurement of the shape and complexity of this cluster provided valuable information over and above EHR-CBC averages. The HD-CBC model more thoroughly explained DeepCBC’s learned signatures yielding reticulocyte characteristics, and hypochromic red cell percentages as additional important features. This broader feature utilisation, unavailable in the conventionally reported blood count parameters, explains the improved detection capability, particularly for early-stage ID before conventional parameters show marked changes. As reported in previous studies, we observed that the strongest signals for ID detection are found in the RET and PLT-F channels of the analyser, but also show that AI analysis of unprocessed flow cytometry data of these channels outperformed individual advanced red cell parameters.^32^

The hard threshold of ferritin = 15 µg*/*L, based upon World Health Organisation (WHO) guidance, presented a challenge for the models, with 60 % of misclassified samples surrounding this boundary. The single threshold value is selected to best discriminate between ID and non-ID states, however there is recognised uncertainty within the 15–30 µg*/*L ferritin range as to where an ideal threshold lies, and any threshold represents a compromise to maximise test sensitivity whilst maintaining specificity.^40, 36^ Indeed, analysis of misclassified samples showed the compromise in selecting a single figure for this purpose and it is notable that for most misclassified samples the true ferritin value fell within the 15–30 µg*/*L range (Fig. 4a).

We acknowledge certain limitations. Current training is limited to predominantly participants of European ancestry with limited exposure to a broader range of ancestries and haemoglobinopathy traits. Other work in the field demonstrates that haemoglobinopathy traits can be effectively discriminated from ID using CBC parameters.^26, 41^ A second limitation is that model training has been restricted to a low-inflammation setting. Published data on the use of individual advanced red cell metrics such as RET-He or Hypo-He, captured within our models, suggest that CBC data are more robust to the effects of inflammation than conventional iron tests such as ferritin.^42^

In summary, our study demonstrates that AI applied to routinely collected CBC data can transform early iron deficiency detection. Where current screening methods identify only 1 in 5 cases of non-anaemic iron deficiency, our approach successfully detects nearly 4 in 5 cases, allowing for proactive iron deficiency management. This advancement offers immediate clinical value without additional testing burden or workflow disruption.

## Acknowledgements

Participants in the INTERVAL, COMPARE and STRIDES studies were recruited with the active collaboration of NHS Blood and Transplant England (www.nhsbt.nhs.uk). The academic coordinating centre at the Department of Public Health and Primary Care at the University of Cambridge received core support from the NIHR Blood and Transplant Research Unit (NIHR203337)*, British Heart Foundation (RG/13/13/30194; RG/18/13/33946) and NIHR Cambridge Biomedical Research Centre (BRC-1215-20014)*, UK Medical Research Council (MR/L003120/1), British Heart Foundation (SP/09/002; RG/13/13/30194; RG/18/13/33946), and by Health Data Research UK, which is funded by the UK Medical Research Council, Engineering and Physical Sciences Research Council, Economic and Social Research Council, Department of Health and Social Care (England), Chief Scientist Office of the Scottish Government Health and Social Care Directorates, Health and Social Care Research and Development Division (Welsh Government), Public Health Agency (Northern Ireland), British Heart Foundation and Wellcome. We thank NIHR BioResource volunteers for their participation and gratefully acknowledge NIHR BioResource centres, NHS Trusts and staff for their contribution. We thank the National Institute for Health and Care Research, NHS Blood and Transplant, and Health Data Research UK as part of the Digital Innovation Hub Programme. The academic coordinating centre would like to thank blood donor centre staff and blood donors for participating. A complete list of the investigators and contributors to the INTERVAL trial is provided in reference [43], COMPARE study in reference [44] and STRIDES trial in reference [45]. *The views expressed are those of the authors and not necessarily those of the NIHR or the Department of Health and Social Care.

## BloodCounts! Consortium

Martijn Schut^25^, Folkert Asselbergs^25^, Sujoy Kar^26^, Suthesh Sivapalaratnam^2,3,24^, Sophie Williams^27^, Mickey Koh^28^, Yvonne Henskens^29^, Bart de Wit^29^, Umberto D’Alessandro^30^, Bubacarr Bah^30^, Ousman Secka^30^, Parashkev Nachev^23^, Rajeev Gupta^31^, Sara Trompeter^31^, Nancy Boeckx^32^, Christine van Laer^32^, Gordon. A. Awandare^33^, Kwabena Sarpong^33^, Lucas Amenga-Etego^33^, Mathie Leers^34^, Mirelle Huijskens^34^, Willem H. Ouwehand^4,12^*^−^*^14^, James HF Rudd^7,9^, Carola-Bibiane Schönlieb^1^, Nicholas S. Gleadall^9,12,14^ and Michael Roberts^1,7^.

^25^ Amsterdam University Medical Centre, Amsterdam, Netherlands. ^26^ Apollo Hospitals, Chennai, India. ^27^ Barts Health NHS Trust, London, United Kingdom. ^28^ Health Services Authority, Singapore. ^29^ Maastricht University Medical Centre, Maastricht, Netherlands. ^30^ MRC The Gambia Unit, Banjul, The Gambia. ^31^ University College London Hospitals, London, United Kingdom. ^32^ University Hospitals Leuven, Leuven, Belgium. ^33^ West African Centre for Cell Biology of Infectious Pathogens, Accra, Ghana. ^34^ Zuyderland Medical Center, Zuyderland, Netherlands.

## STRIDES NIHR BioResource

John R Bradley^5^*^−^*^8^, Nathalie Kingston^6,12^, Kathleen Stirrups^6,12^, Paul Townsend^6^, Jacinta Lee^6^, Nigel Ovington^6^, Hannah Stark^6^.

## STRIDES Trial

For authors and affiliations, see reference [45]. A finalised list of authors and affiliations will be printed here before acceptance of the manuscript.

## Declarations

### Ethics approval and consent to participate

The INTERVAL, COMPARE, and STRIDES trials are compiled into the Blood Donors Studies BioResource (BDSB) with Research Ethics Committee (REC) reference 20/EE/0115.

### Consent for publication

All authors have reviewed and approved the submission.

### Data availability

Access to BDSB data can be applied for from the Blood Donors Studies Data Access Committee.

### Materials availability

Not applicable.

### Code availablity

Models and code are available upon request and will be made publicly available upon acceptance of the manuscript.

### Author contributions

D.K. and J.T. performed the analysis and drafted the manuscript. D.K. performed data pre-processing and evaluation for all models, and trained the EHR-CBC and HD-CBC models. J.T. conceived the study, J.T., W.H.O and M.R. designed the study. S.D. designed and trained the DeepCBC model. M.B., J.G., S.K., W.H.O., O.S., D.S., D.V., J.R., E.D.A., C.-B.S., P.N., M.R. consulted throughout the work on methodology, design and writing. J.R.B., N.K., and K.S. (NIHR BioResource), and E.D.A., S.K., W.H.O., and D.R. (Donor Health Studies) were instrumental in the conception, design, and execution of the INTERVAL, COMPARE, and STRIDES trials on which this study is built upon. S.S., N.S.G., and M.R. received the funding for the study. M.R. provided supervision of the researchers, M.R. and W.H.O provided edits to the early manuscript and subsequent versions. All authors approved the final manuscript.

## 4 Methods

### 4.1 Data sources

We utilised data from three cohort studies within England blood donor populations: INTERVAL^43^ (ISRCTN24760606), COMPARE^44^ (ISRCTN90871183), and STRIDES^45^ (ISRCTN10412338). We used rich complete blood count (R-CBC) data from all three trials for pre-training of the DeepCBC model. Results are quoted for the INTERVAL dataset, due to the availability of paired CBC and iron homeostasis test results.

INTERVAL^43^ was an randomised controlled trial (RCT) assessing the safety of different blood donation frequencies (8, 10, 12 weeks for males; 12, 14, 16 weeks for females) over a 24-month period (see Fig. 7a), with a subgroup also monitored up to 48 months.^46^ This cohort is representative of a healthy population, where iron deficiency (ID) is the primary cause of anaemia, making it very suitable for ID detection model development. In addition the Sysmex CBC measurements and extensive iron homeostasis testing (ferritin, transferrin, serum iron, soluble transferrin receptor, hepcidin), the dataset also consists of C-reactive protein (CRP) levels and various subject characteristics such as sex, age, weight, and height. The inflammation burden in this population was low, with an average CRP of 1.97 mg*/*L (92 % of participants with CRP under 5 mg*/*L), and an average white blood cell count (WBC) of 6.95 *×* 10^9^ cells*/*L (95 % of participants with WBC under 10 *×* 10^9^ cells*/*L). Data is available for 48,725 subjects and after pre-processing (such as excluding subjects with missing or unreliable CBC or ferritin measurements, or unreliable subject information), we used data from 46,289 participants for analysis (Fig. 7b).

**Fig. 7:**
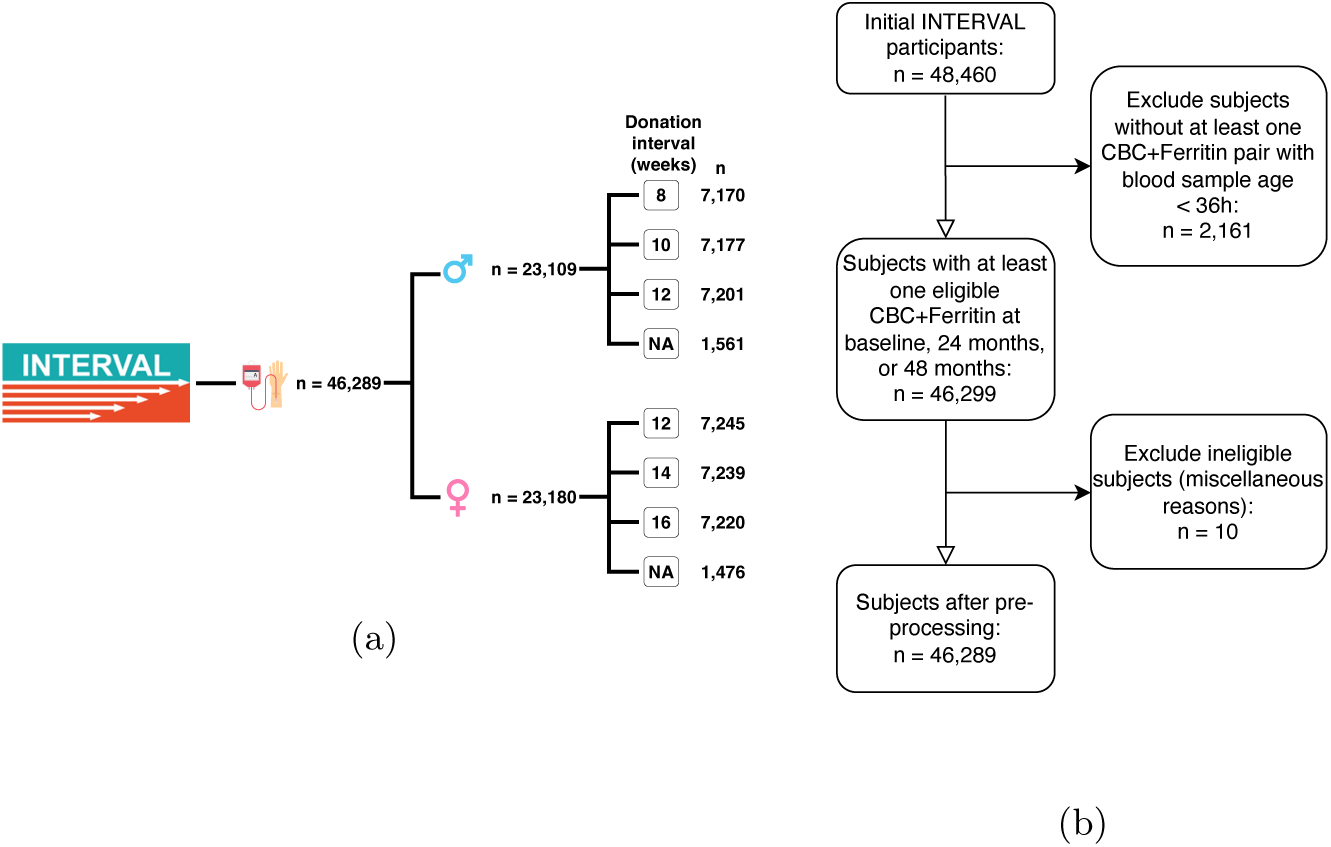
INTERVAL blood donation trial data and participant pre-processing. **a,** INTERVAL trial design, blood donors were split into trial arms with varying donation frequencies in weeks. Participants who had enrolled at baseline but did not complete their baseline questionnaire were not invited to further study donations (denoted as “NA” in the figure). **b,** Sample selection flowchart of pre-processing steps for the INTERVAL trial data.

We utilise COMPARE and STRIDES for pre-training DeepCBC, using their combined R-CBC data. For COMPARE,^44^ we use 39,093 R-CBCs from 29,021 participants. For STRIDES,^45^ we use 61,660 R-CBCs from 83,861 participants. Unlike INTERVAL, we did not perform filtering or pre-processing on the COMPARE and STRIDES data as their purpose was simply to enable DeepCBC’s variational autoencoder (VAE) component to learn characteristics of R-CBC data in an unsupervised way.

### 4.2 Clinical thresholds for ID, anaemia, and donation

For this study, ID was defined as ferritin *<* 15 µg*/*L, following the World Health Organisation (WHO) recommended thresholds.^36^ Low iron stores were defined as a ferritin value within the range of 15–30 µg*/*L following the approach taken in the the FIND’EM study and thresholds adopted in clinical practice.^35, 18^ Subject sex was not used when defining the lower limit of ferritin, following WHO guidance.^36, 47^ Anaemia was defined as a haemoglobin level of under 130 g*/*L for males and 120 g*/*L for females, again following WHO guidance.^37^

During the INTERVAL trial regular NHS Blood and Transplant (NHSBT) donation deferral strategies were in place at the time. Subject haemoglobin levels were estimated using the gravimetric copper sulphate test at the time of donation. Haemoglobin for subjects who failed the copper sulphate test was re-measured using the spectrophotometric *Hemocue* test. Subjects with haemoglobin levels in the ranges 125–134 g*/*L and 115–124 g*/*L, for males and females respectively, were deferred for a period of three months, while subjects with lower haemoglobin levels were deferred for a period of 12 months. After deferral, the donors rejoined the trial. Donors with three consecutive low haemoglobin results on the Hemocue test were withdrawn from blood donation altogether.^43^

To model correlations between CBC parameters and ferritin, penalised cubic spline models with 10 knots for ferritin were produced using the pyGAM^48^ package (see Fig. 3b & 3c).

### 4.3 CBC data preparation

CBC measurements for INTERVAL, COMPARE, and STRIDES were recorded on Sysmex XN haematology analysers. Raw measurement data from CBCs were retained. In this work, we trained artificial intelligence (AI) models on various stages of the CBC measurement pathway. During CBC analysis, blood samples are processed by the Sysmex XN analysers across several measurement channels: WNR, WDF, RET, PLT-F (single-cell flow cytometry), RBC and PLT (hydrodynamically focused DC), and SLS haemoglobin (Fig. 1). From these channels, the analysers calculate CBC parameters, and Sysmex Interpretive Program (IP) messages flag potential abnormalities with “Q-Flag” fields indicating suspected abnormality levels. For our analyses, we categorised CBC data into three types: **EHR-CBC** (standard parameters commonly reported in Electronic Health Records), **High-Dimensional CBC (HD-CBC)** (comprising intermediate-rich CBC data such as extended summary CBC reports, instrument messages and quality flags, combined with engineered features extracted from raw single-cell measurements), and **Rich CBC (R-CBC)** (the unprocessed single-cell measurement data). Raw Sysmex data were not recorded for all samples in the trial. In this work, we use the available extended data only when specified; otherwise, we use the full dataset where only EHR-CBCs were recorded.

EHR-CBC parameters referred to in this work include white blood cell count (WBC), red blood cell count (RBC), haemoglobin concentration, haematocrit (HCT), mean corpuscular volume (MCV), mean corpuscular haemoglobin (MCH), MCH concentration (MCHC), platelet count (PLT), mean platelet volume (MPV), platelet distribution width (PDW), red cell distribution width (RDW), neutrophil count (NEUT), lymphocyte count (LYMPH), monocyte count (MONO), eosinophil count (EO), basophil count (BASO), nucleated red cell count (NRBC), and immature granulocyte count (IG).

To maintain the clinical reliability of our dataset, we adhered to exclusion criteria similar to those used in hospital haematology laboratories. Samples with suspected red cell turbidity, red cell agglutination, or platelet clumping, based on IP messages, were excluded, as such conditions are known to produce unreliable measurements. Additionally, samples were excluded if multiple clinically implausible metrics were observed concurrently, as these are indicative of sample clotting or other issues. Specifically, we removed samples with a platelet count below 50,000*/*µL, a white blood cell count below 1,000*/*µL, and a haematocrit under 20 %, when these occurred simultaneously.

Often, due to issues with the sample or CBC instrument, a repeat measurement may be performed. In such cases, we compared the first two measurements for each sample to determine which should be retained. Since information about the laboratory procedures during the trials was not available, we compared the EHR-CBC variables between measurements. If the two measurements agreed within one standard deviation on all but one variable, the first measurement was retained, as there is no reason to doubt its validity. If this criterion was not met, all measurements for that sample were discarded, as there was no robust method to reconcile discrepancies between measurements.

#### 4.3.1 R-CBC data

Fig. 2a illustrates how R-CBC data is generated, including the hydrodynamically focussed DC (direct current) method to count red blood cells (RBC) and platelets (PLT), shown in red, and fluorescence flow cytometry for white cell differential measurements (WNR & WDF channels), advanced optical RBC, reticulocyte and PLT counts (RET channel), and closer platelet inspection, especially focussing on immature platelets (PLT-F channel). The data is saved as 1D histograms (hydrodynamically focused DC) and 2D (RET) to 4D (WDF, WNR, PLT-F) flow cytometry scattergrams.

#### 4.3.2 HD-CBC data

To offer a more interpretable alternative to the R-CBC model, we constructed HD-CBC data out of intermediate-rich CBC data such as extended summary CBC reports, instrument messages and quality flags, and manually derived features from the R-CBC, increasing the number of features to 716, while maintaining interpretability. IP messages and Q-Flags were encoded as binary indicators (0/1) to capture abnormality presence, and categorical message states (e.g., “DISCRETE” or “ERROR”) were encoded into individual columns. Specific measurement abnormalities (e.g., “haemoglobin out of measurement range” or “unreliable RBC”) were likewise encoded as distinct fields. Some Q-Flags are given as a continuous score by the analyser, which was retained. To allow for more interpretability of feature importances, features with *>* 0.8 correlation to any of the EHR-CBC features were removed.

##### Processing hydrodynamically focused DC measurements as features for the HD-CBC

Sysmex XN-2000 haematology analysers record blood cell measurements using multiple measurement channels. Various red cell and platelet characteristics are measured using a hydrodynamically focused DC (direct current) detection method. The resulting histogram is recorded as 128 separate measured points (Fig. 8).

**Fig. 8:**
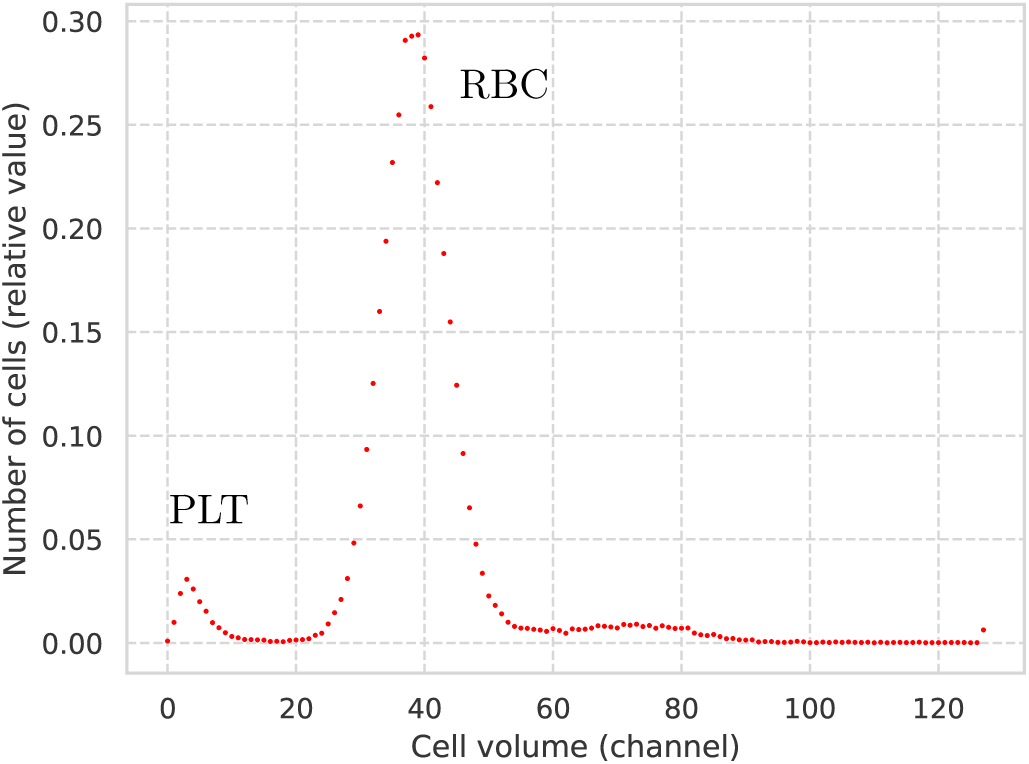
Example histogram of 1D hydrodynamically focused DC data, with peaks for the PLT (left) and RBC (right).

To transform this 128-point measurement into meaningful features for the HD-CBC, we used a 1-dimensional convolutional neural network (CNN) autoencoder to learn lower-dimensional embeddings (effectively compressions) of the data. The employed architecture is displayed in Fig. 9.

**Fig. 9:**
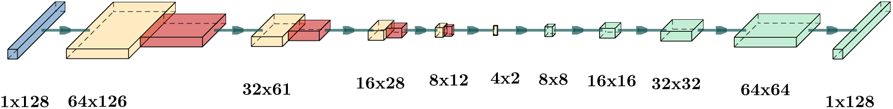
Architecture of employed 1D-CNN autoencoder for compression of the hydrodynamically focused DC method data. The input (blue) is a single-channel 128-point measurement that is propagated through a series of convolution (yellow) and max-pooling (red) blocks to the embedding space, creating a 4 *×* 2 signal. The embedding is subsequently upsampled by transposed convolutions (green) to reproduce the input signal. The data dimensions at each layer are annotated below the layers.

Unsupervised model training was conducted using the Adam optimiser for 200 epochs with a batch size of 1024. A train-test split in the INTERVAL data of 90 %:10 % was used to train and evaluate the model quality. The trained model was then used to generate 8-dimensional embedding vectors of all hydrodynamically focused DC method measurements. These 8-dim vectors were appended to their respective HD-CBCs for the analysis in this work.

##### Processing flow cytometry measurements as features for the HD-CBC

We also augment the HD-CBC with derived features from the WNR, WDF, RET and PLT-F flow cytometry measurement channels of the Sysmex XN-2000 analysers. The flow cytometry data includes side-fluorescence light (SFL), forward-scattered light (FSC), side-scattered light (SSC), and forward-scattered light width (FSCW) measurements of cells in the blood sample. WNR, WDF, and PLT-F channels use all measurements, while the RET channel only contains SFL and FSC. Each sample contained approximately 30,000–90,000 individual cell measurements.

To derive meaningful features from these data, we chose to fit multivariate Gaussian distributions to the various cell populations in the scattergrams and record their mean vectors and covariance matrices as features for the HD-CBC. To achieve this, we first removed saturated events (i.e., measurements beyond the detection range), then randomly sampled 1,000,000 single-cell events from the whole INTERVAL dataset, resulting in a scattergram representative of the study population average. We then manually created polygon gatings for each Sysmex measurement channel, using Sysmex documentation images. These gates were designed to capture the characteristic distribution of each cell population rather than to achieve perfect classification of every cell, as our objective was to extract representative distribution parameters rather than precise cell counts. Nevertheless, comparing across individual samples, we have found consistency in cell population discrimination using our gatings. We subsequently used the gatings on each individual sample to fit the multivariate Gaussian distributions.

We found the multivariate Gaussian distributions reflect the distinct cell populations well (Fig. 10). Gaussians were fitted to the flow cytometry data for WNR, WDF, and PLT-F in 4D, whilst the RET channel distributions were fit in 2D. This approach enabled us to leverage the flow cytometry data as part of the HD-CBC, comprising 185 of the 716 features.

**Fig. 10:**
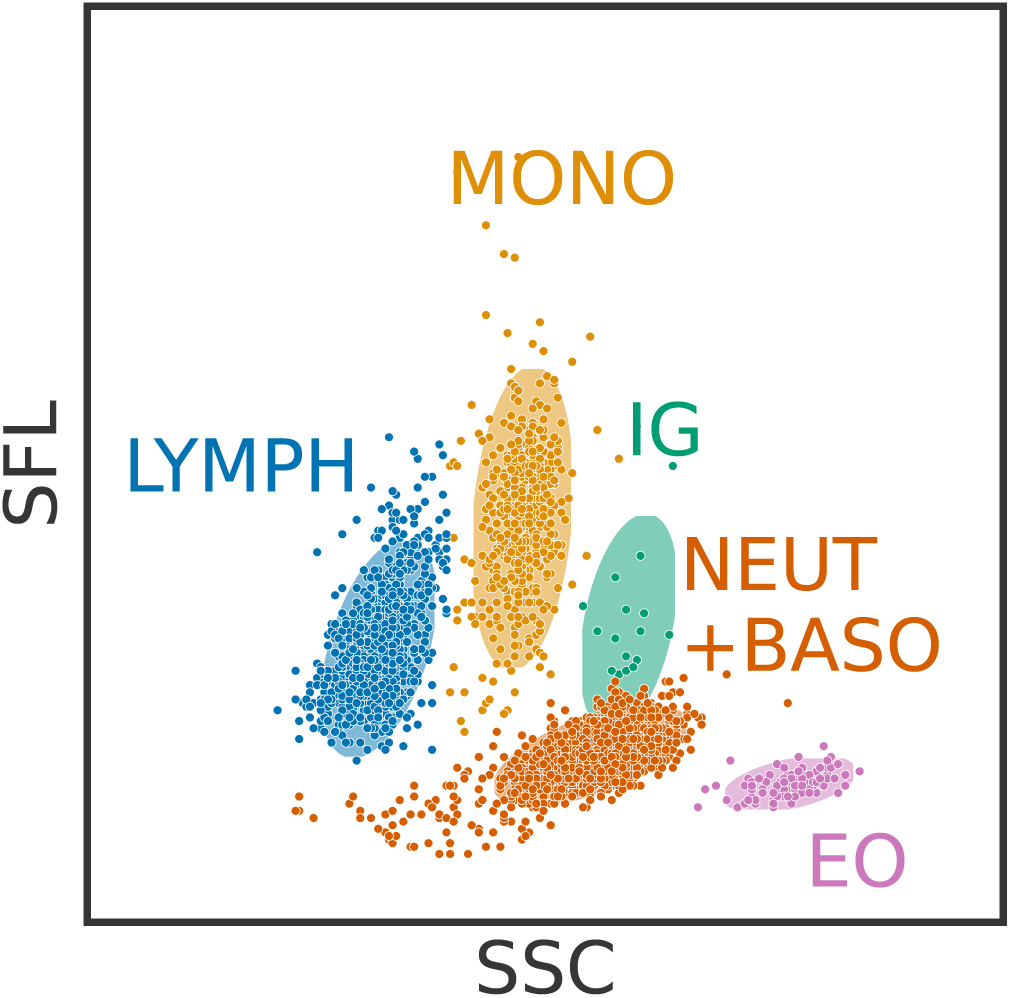
Sysmex WDF channel measurement for a sample in the INTERVAL trial using the Sysmex XN-2000 analyser. Only SFL and SSC flow cytometry channels are shown. Cells are coloured based on their type (as determined by manual gating), and ellipses illustrate the multivariate Gaussian distributions fit to each cell population. LYMPH, lymphocytes; MONO, monocytes; IG, immature granulocytes; NEUT, neutrophils; BASO, basophils; EO, eosinophils.

#### 4.3.3 CBC data adjustment for technical covariates

In order to remove variance explained by technical or environmental effects, we followed a similar feature adjustment procedure to Astle et al.^30^ We adjusted for four distribution-shifting effects: delay between sample acquisition and analysis, time since the beginning of the study, time of day at sample analysis, and day of the week at sample analysis. These effects were modelled using cubic spline generalised additive models (GAMs) for all HD-CBC and EHR-CBC features separately for each analyser. As Astle et al^30^ have done, we adjusted the positively supported indices (e.g. cell counts) on the log-scale and the proportion-supported indices (e.g. ratios or percentages) on the logit scale. The GAM models predicted the expected value of a blood feature given only information on the effects outlined above. Including an intercept term, each term of the model thus effectively models the expected change to the mean of the considered blood feature only due to a specific technical effect. Each feature is then adjusted by the difference between the GAM model’s prediction and the mean of the feature on the adjustment scale. An example of this is shown in Fig. 11, where the white blood cell count is plotted over the delay between sample acquisition and analysis is shown before and after the adjustment.

**Fig. 11:**
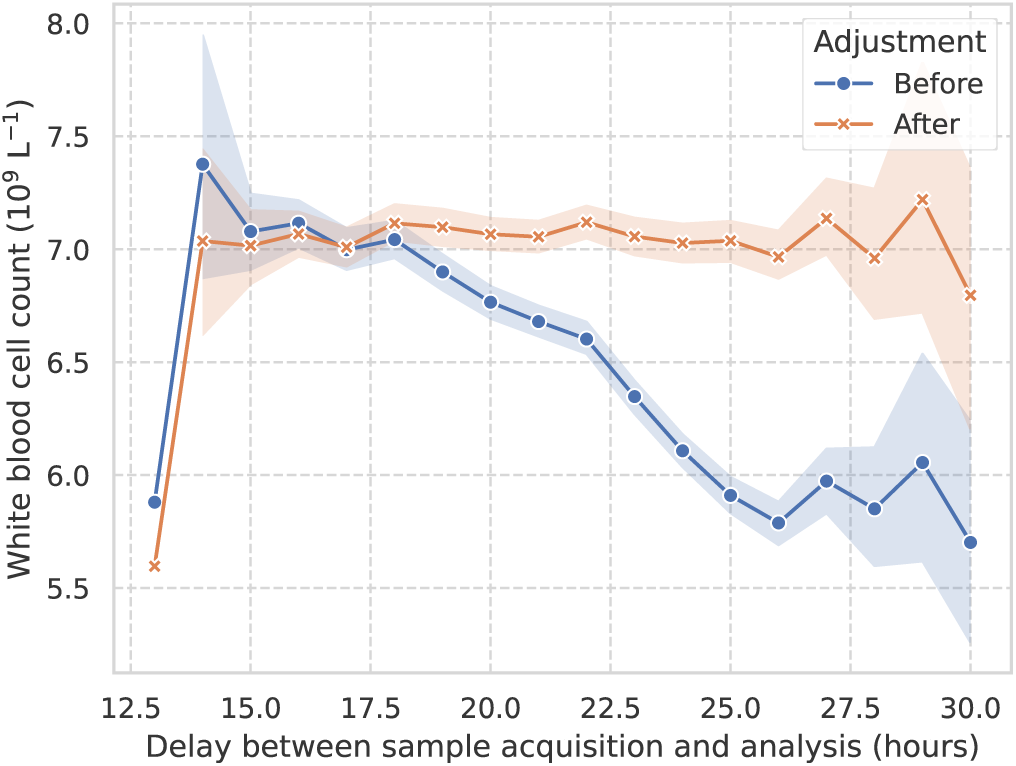
White blood cell count changes with venepuncture-to-analysis delay. Mean WBC measured on a single Sysmex instrument during one-hour intervals. Shaded regions interpolate the 95 % confidence intervals of the means. Blue: unadjusted data; orange: covariate-adjusted data demonstrating elimination of delay-dependent measurement bias.

### 4.4 Statistical tests

Where p-values have been reported, these were derived using the independent Student’s *t*-test for normally distributed variables and Mann–Whitney *U* test for non-normally distributed variables. Satisfaction of the normality assumption was ascertained using the Shapiro-Wilk test and asserting a p-value of *<* 0.001.

### 4.5 Artificial intelligence models

In this study, we employed multiple machine learning models. For tabular data (EHR-CBC and HD-CBC), we used XGBoost or Multilayer-Perceptron (MLP) models. For the R-CBC data, we used an end-to-end Deep Learning-based variational autoencoder (VAE) approach.

All time points from INTERVAL were used for model training. As such, samples from the same donor could appear up to three times in the dataset (baseline, 24 months, and 48 months). The INTERVAL data was split into training (69,821 samples), validation (7,568 samples), and test (8,951 samples) sets, with stratification by donors to prevent overlap across splits. Model training was performed on the training set, hyperparameters, calibration and operating thresholds were optimised on the validation set, and reported evaluations were conducted on the test set. The classification threshold was determined as the point maximising sensitivity and specificity on the validation receiver operating characteristic (ROC) curve.

All ID detection models underwent probability calibration with logistic regression.^49^ The validation dataset was used to calibrate the classifiers.

#### 4.5.1 XGBoost models

We utilised XGBoost^20^ (eXtreme Gradient Boosting) classifiers for the EHR-CBC and HD-CBC ID detection models. XGBoost was selected due to its robust performance on tabular data and its inherent interpretability. The hyperparameter configuration was determined using a Bayesian optimisation over multiple configurations in Weights & Biases,^50^ using 5-fold cross validation.

XGBoost models were trained in less than 1 minute on 188 Intel Xeon CPUs in parallel.

#### 4.5.2 Deep learning models

To fully exploit the richness of the R-CBC data, we developed a deep learning approach named ‘DeepCBC’. DeepCBC can directly utilise the high-dimensional flow cytometry and impedance signals from the Sysmex XN analyser. The DeepCBC architecture first encodes the complex R-CBC data into a lower-dimensional latent representation through a variational autoencoder (VAE). Each measurement channel in the R-CBC data was processed separately: from the 4D Sysmex channels (WNR, WDF, PLT-F), the FSC, SSC, and SFL flow cytometry channels were reduced to three 2D images each by taking maximum intensity projections along the third axis, while the RET channel, inherently 2D, was processed using two distinct views (FSC vs. SFL*×*2 and log-FSC vs. SFL). These 2D representations were then transformed into 128-dimensional vectors using convolutional neural networks (CNNs; EfficientNet-B0 architecture^51^). Similarly, the 1D impedance signals (RBC and PLT channels, each with 128 measurement points) and all FSCW measurements (WNR, WDF, PLT-F) were separately encoded using 1-dimensional CNNs into 16-dimensional vectors. All resulting representation vectors were concatenated and fed through a multilayer perceptron (MLP) encoder to form a unified 256-dimensional latent space representation. The data was then reconstructed by symmetric CNN decoders, enabling unsupervised training of the VAE. The VAE pre-training was performed on combined training data from the INTERVAL, COMPARE, and STRIDES studies, allowing the model to learn robust initial weights and general features in an unsupervised manner. Subsequently, an MLP classification head was attached to the end of the encoder to act as the classifier for the classification tasks: ferritin *<* 15 µg*/*L, sex, presence of O blood type, and RhD type. For the ferritin task, a linear classification head (single-neuron output) was attached to the encoder to predict ID status (ferritin *<* 15 µg*/*L). The whole model was then fine-tuned in two stages using only the INTERVAL training data: initially by freezing the encoder and optimising only the linear head, followed by full fine-tuning of all layers. To address class imbalance, samples with ferritin below the threshold were oversampled by a factor of 2.5 during training. The encoder was frozen for the non-disease tasks, and a 2-hidden layer MLP classification head was attached to the encoder (dimensions: 256 *→* 512 *→* 128 *→* 1). For these tasks, the encoder was effectively used as a dimensionality reduction tool, and the MLP was trained on the 256-dimensional latent space representations of the R-CBC data. The final DeepCBC model contained approximately 91 million trainable parameters. The best-performing model on the validation set was selected and subsequently evaluated on the independent test set.

#### 4.5.3 Model explainability

For EHR-CBC and HD-CBC XGBoost models detecting ID, we used the Shapley Additive Explanations (SHAP)^52, 53^ Python package to calculate and plot model feature importances and single-sample decisions. In particular, we used SHAP’s TreeExplainer^54^ to estimated model Shapley values (using a random subset of 1000 training examples as the background distribution for the estimation).

For DeepCBC, we can leverage the VAE architecture to perform ‘class difference interpolation’. We first calculate the semantic direction in the model latent space between mean latent feature vectors of each class, and subsequently move along this direction from the latent space representation of a real sample. During this process, we can regularly generate a sample in data space using the model’s decoder, allowing us to visualise smooth transitions between data from one class moving to different classes. For parameters with continuous information, like ferritin, we used samples below the 10th and above the 90th percentiles within our training data as examples of the two classes from which to calculate the semantic direction vector. Within the Results section (Fig. 2c and 5c), we display multiple class difference interpolation scattergrams where density is lost in the blue regions and gained in the red ones when traversing from the negative to the positive class.

## Appendix A Extended Data

### A.1 Confusion matrices for sex, presence of blood type O, and RhD type classification

**Fig. A1:**
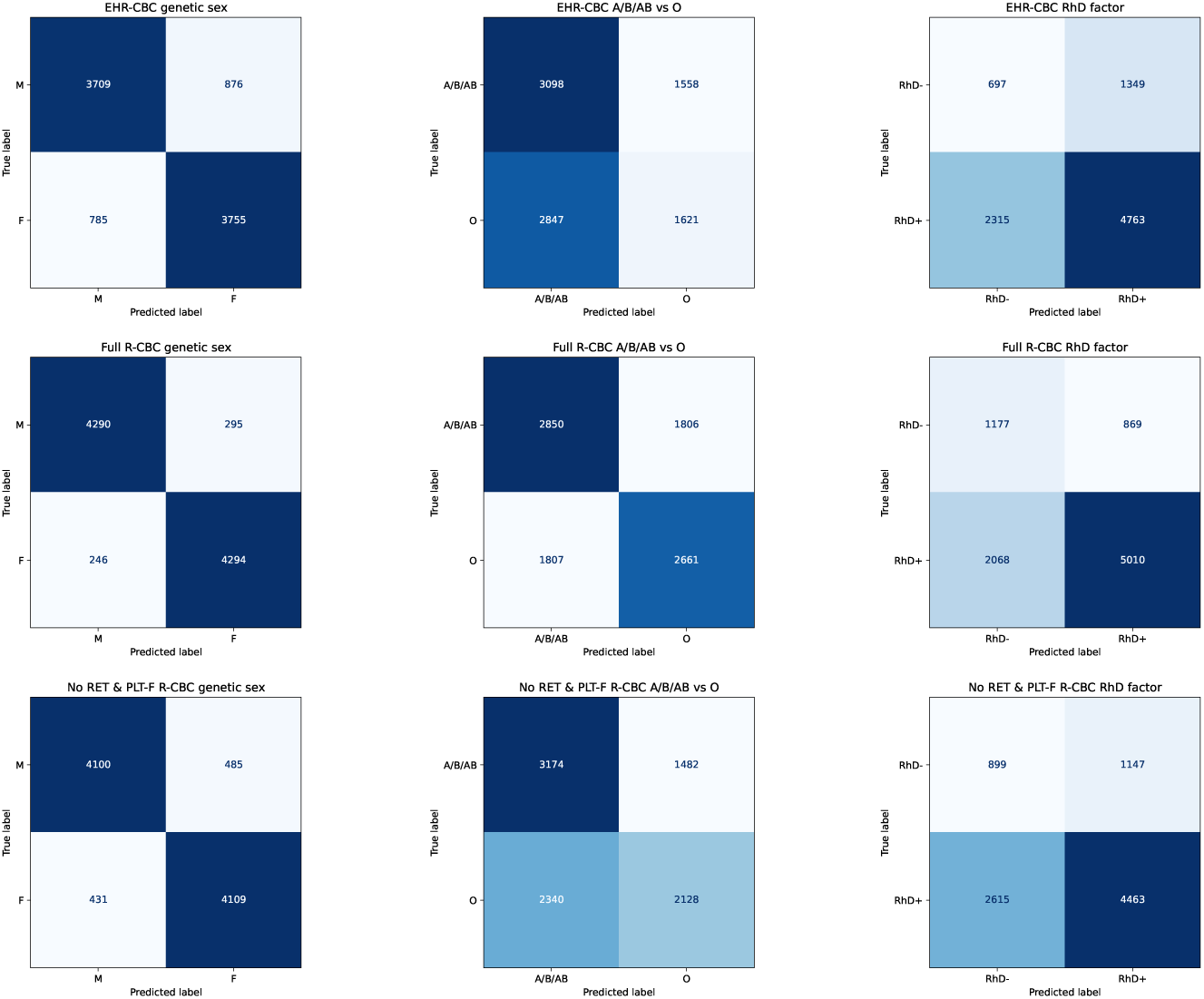
Confusion matrices for the classification tasks from section 2.1, evaluated on the held-out test set from the INTERVAL trial. Numbers of samples vary between tasks as not every blood sample had all three labels available (sex, ABO blood type, RhD type).

### A.2 Iron status in INTERVAL

**Table A1:** INTERVAL iron status by sex and time point.

| Time point | Sex | Iron replete | Low stores | Iron deficient |  |
| --- | --- | --- | --- | --- | --- |
|  |  |  |  | Non-anaemic | Anaemic |
| Baseline | Males | 15,482 (70.8 %) | 4,522 (20.7 %) | 1,677 (7.7 %) | 188 (0.9 %) |
|  | Females | 9,415 (43.1 %) | 7,019 (32.1 %) | 4,670 (21.4 %) | 747 (3.4 %) |
| 24 months | Males | 7,206 (52.7 %) | 3,874 (28.3 %) | 1,734 (12.7 %) | 864 (6.3 %) |
|  | Females | 5,444 (41.7 %) | 4,003 (30.6 %) | 2,451 (18.8 %) | 1,165 (8.9 %) |

**Fig. A2:**
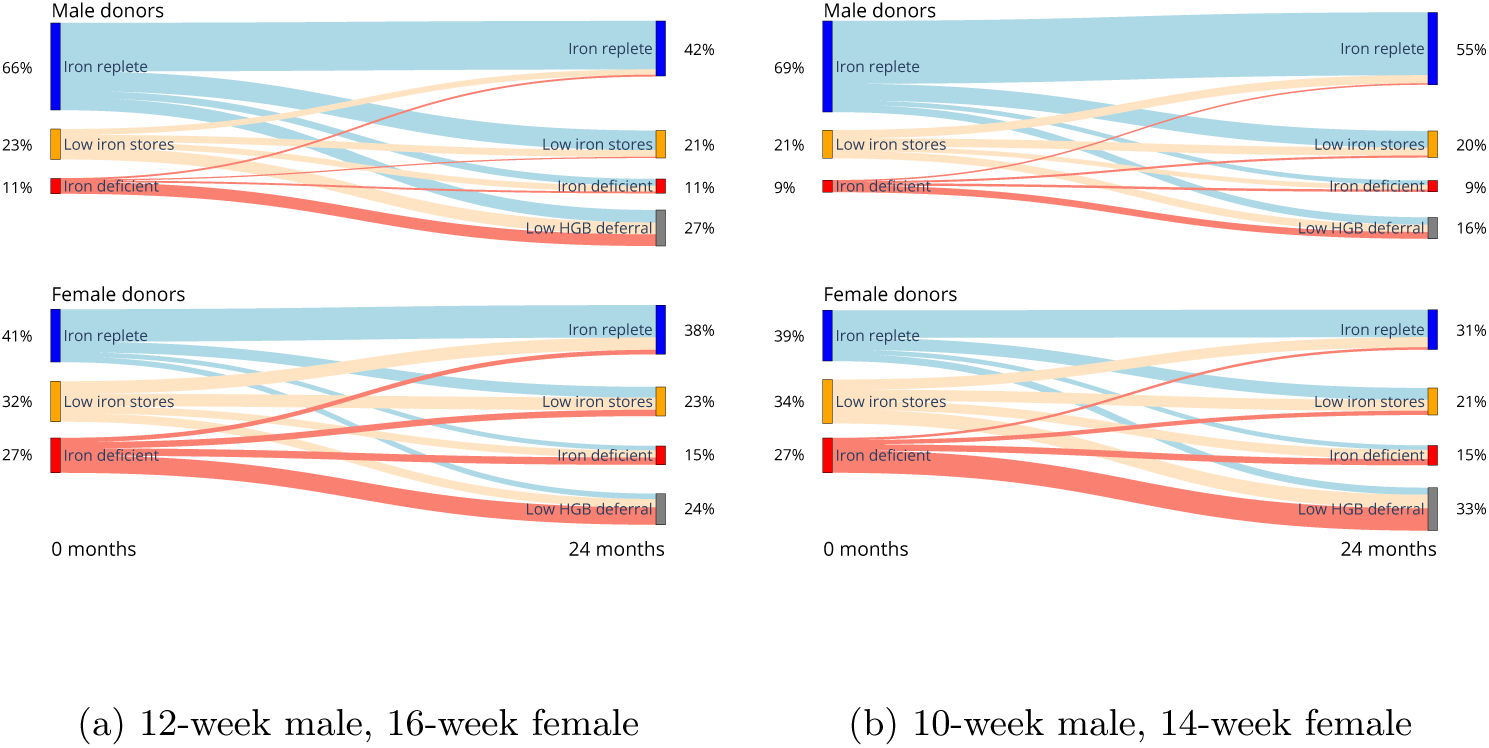
Sankey plot of shift in subject iron status over PHASE 1 of INTERVAL for different trial arms. Subjects with at least one low-haemoglobin donation deferral separated.

**Table A2:** INTERVAL baseline EHR-CBC parameters by sex and iron status, together with statistical test evaluation.

| Feature | Female |  |  | Female comparison |  |  | Male |  |  | Male comparison |
| --- | --- | --- | --- | --- | --- | --- | --- | --- | --- | --- |
|  | ID | IR | t-stat | p-value | Cohen's d | ID | IR | t-stat | p-value | Cohen's d |
| Haemoglobin (g/L) | 128.91 ± 8.82 | 136.24 ± 8.75 | 50.187 | <0.001 | 0.835 | 141.21 ± 9.10 | 150.76 ± 9.67 | 42.288 | <0.001 | 0.995 |
| MCV (fL) | 89.74 ± 5.01 | 93.95 ± 4.43 | 54.741 | <0.001 | 0.904 | 88.14 ± 5.03 | 92.17 ± 4.37 | 38.792 | <0.001 | 0.906 |
| MCH (pg) | 28.02 ± 1.89 | 30.07 ± 1.58 | 72.583 | <0.001 | 1.199 | 27.78 ± 1.92 | 30.04 ± 1.56 | 60.124 | <0.001 | 1.404 |
| MCHC (g/dL) | 31.27 ± 1.07 | 32.07 ± 1.08 | 45.008 | <0.001 | 0.743 | 31.56 ± 1.09 | 32.65 ± 1.11 | 42.450 | <0.001 | 0.991 |
| RDW (fL) | 47.95 ± 4.41 | 45.28 ± 3.45 | -42.015 | <0.001 | -0.694 | 46.31 ± 3.98 | 44.02 ± 3.42 | -28.109 | <0.001 | -0.657 |
| PLT (billion/L) | 260.72 ± 69.54 | 252.05 ± 62.04 | -8.073 | <0.001 | -0.133 | 229.59 ± 60.85 | 223.58 ± 53.92 | -4.702 | <0.001 | -0.110 |
| WBC (billion/L) | 6.97 ± 1.72 | 7.09 ± 1.79 | 3.925 | <0.001 | 0.065 | 6.22 ± 1.59 | 6.46 ± 1.64 | 6.316 | <0.001 | 0.148 |
| NEUT (billion/L) | 4.00 ± 1.32 | 4.03 ± 1.34 | 1.252 | 0.210 | 0.021 | 3.52 ± 1.20 | 3.63 ± 1.17 | 3.851 | <0.001 | 0.090 |
| LYMPH (billion/L) | 2.18 ± 0.61 | 2.26 ± 0.65 | 7.998 | <0.001 | 0.132 | 1.89 ± 0.64 | 2.02 ± 0.78 | 6.842 | <0.001 | 0.160 |
| MONO (billion/L) | 0.56 ± 0.16 | 0.56 ± 0.16 | -0.917 | 0.359 | -0.015 | 0.57 ± 0.16 | 0.58 ± 0.17 | 3.009 | 0.003 | 0.070 |
| EO (billion/L) | 0.16 ± 0.13 | 0.17 ± 0.14 | 2.208 | 0.027 | 0.036 | 0.18 ± 0.13 | 0.18 ± 0.14 | 0.340 | 0.734 | 0.008 |
| BASO (billion/L) | 0.05 ± 0.02 | 0.05 ± 0.02 | -1.301 | 0.193 | -0.021 | 0.05 ± 0.02 | 0.05 ± 0.02 | -2.089 | 0.037 | -0.049 |
| NRBC (billion/L) | 0.00 ± 0.01 | 0.01 ± 0.02 | 4.298 | <0.001 | 0.071 | 0.01 ± 0.02 | 0.01 ± 0.02 | 2.520 | 0.012 | 0.059 |
| IG (billion/L) | 0.03 ± 0.01 | 0.03 ± 0.02 | 5.899 | <0.001 | 0.097 | 0.02 ± 0.03 | 0.03 ± 0.02 | 6.861 | <0.001 | 0.160 |

Values are presented as mean ± standard deviation. ID = Iron deficient (ferritin < 15 µg/L), IR = Iron replete (ferritin > 30 µg/L). Statistical comparisons use independent *t*-tests between iron replete and iron deficient groups within each sex. Cohen's *d* represents standardised effect size.

### A.3 ID detection performance per model

**Fig. A3:**
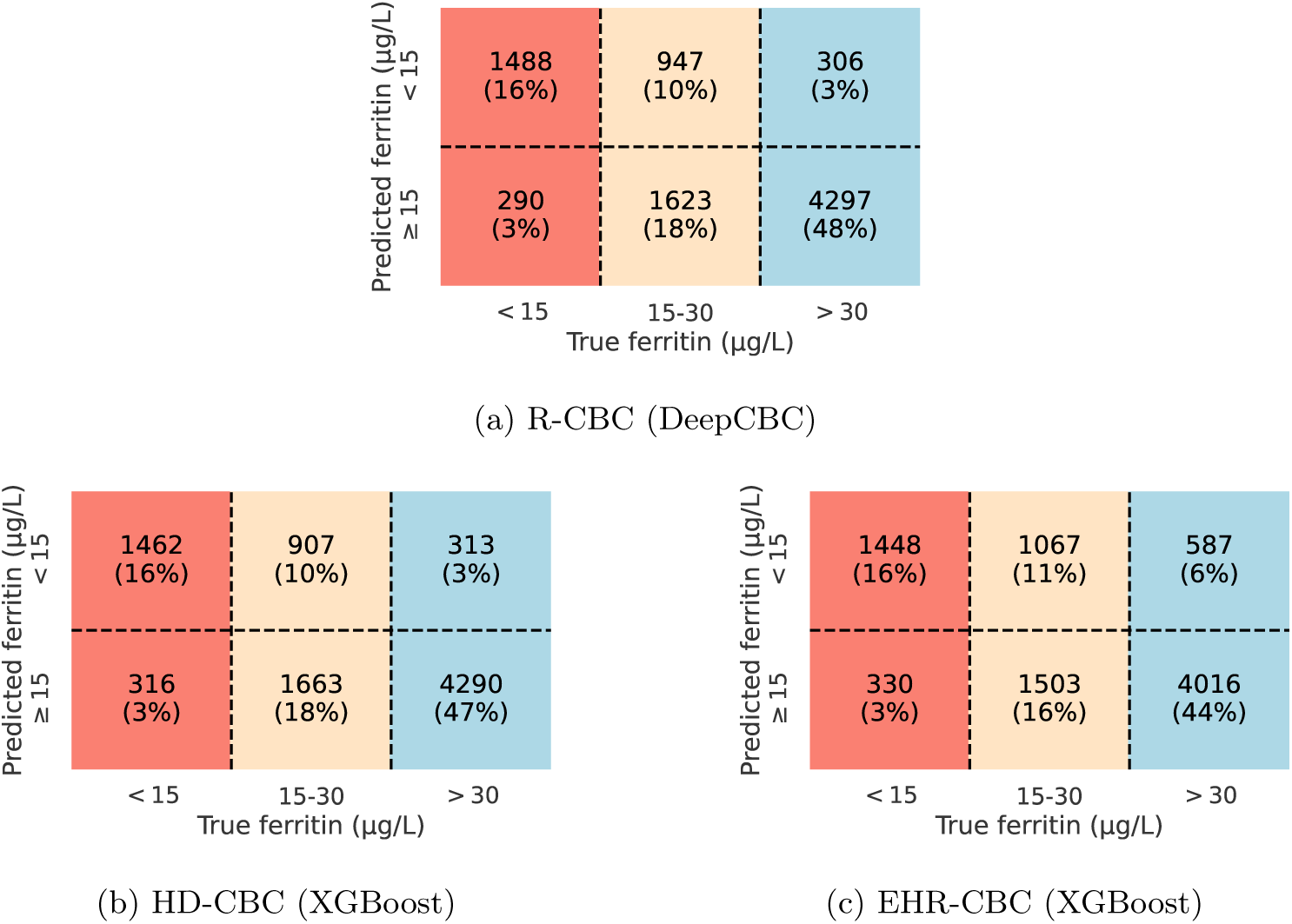
Confusion matrices for ID detection on the held out test set for the three models studied. The confusion matrix is separated into ID, low iron stores, and iron replete regions.

**Table A3:** Model performance metrics by trial arm ([Sex][Donation interval in weeks]). ROC AUC, receiver operator characteristic area under the curve; AP, average precision; PPV, positive predictive value.

| Model | Trial Arm |  |  |  |  |  |  |
| --- | --- | --- | --- | --- | --- | --- | --- |
|  | M08 | M10 | M12 | F12 | F14 | F16 | All |
| **ROC AUC** |  |  |  |  |  |  |  |
| EHR-CBC (XGBoost) | 0.891 | 0.870 | 0.879 | 0.850 | 0.843 | 0.854 | 0.870 |
| HD-CBC (XGBoost) | 0.925 | 0.910 | 0.914 | 0.890 | 0.891 | 0.893 | 0.908 |
| R-CBC (DeepCBC) | 0.933 | 0.920 | 0.911 | 0.886 | 0.893 | 0.893 | 0.909 |
| **AP** |  |  |  |  |  |  |  |
| EHR-CBC (XGBoost) | 0.653 | 0.626 | 0.576 | 0.673 | 0.704 | 0.669 | 0.659 |
| HD-CBC (XGBoost) | 0.742 | 0.709 | 0.631 | 0.749 | 0.789 | 0.745 | 0.741 |
| R-CBC (DeepCBC) | 0.767 | 0.724 | 0.611 | 0.759 | 0.793 | 0.739 | 0.747 |
| **Balanced Accuracy** |  |  |  |  |  |  |  |
| EHR-CBC (XGBoost) | 0.812 | 0.796 | 0.818 | 0.765 | 0.756 | 0.764 | 0.791 |
| HD-CBC (XGBoost) | 0.848 | 0.820 | 0.837 | 0.815 | 0.799 | 0.810 | 0.826 |
| R-CBC (DeepCBC) | 0.846 | 0.841 | 0.838 | 0.804 | 0.804 | 0.828 | 0.831 |
| **Sensitivity** |  |  |  |  |  |  |  |
| EHR-CBC (XGBoost) | 0.804 | 0.768 | 0.785 | 0.827 | 0.838 | 0.827 | 0.813 |
| HD-CBC (XGBoost) | 0.850 | 0.790 | 0.785 | 0.843 | 0.832 | 0.806 | 0.821 |
| R-CBC (DeepCBC) | 0.850 | 0.839 | 0.785 | 0.832 | 0.855 | 0.840 | 0.837 |
| **Specificity** |  |  |  |  |  |  |  |
| EHR-CBC (XGBoost) | 0.820 | 0.823 | 0.850 | 0.703 | 0.674 | 0.702 | 0.770 |
| HD-CBC (XGBoost) | 0.845 | 0.851 | 0.889 | 0.787 | 0.766 | 0.815 | 0.830 |
| R-CBC (DeepCBC) | 0.843 | 0.844 | 0.890 | 0.776 | 0.753 | 0.816 | 0.825 |
| **PPV** |  |  |  |  |  |  |  |
| EHR-CBC (XGBoost) | 0.496 | 0.446 | 0.397 | 0.494 | 0.485 | 0.446 | 0.466 |
| HD-CBC (XGBoost) | 0.547 | 0.494 | 0.470 | 0.582 | 0.566 | 0.558 | 0.544 |
| R-CBC (DeepCBC) | 0.544 | 0.497 | 0.475 | 0.567 | 0.560 | 0.570 | 0.543 |
| **F1** |  |  |  |  |  |  |  |
| EHR-CBC (XGBoost) | 0.614 | 0.564 | 0.527 | 0.619 | 0.615 | 0.579 | 0.592 |
| HD-CBC (XGBoost) | 0.666 | 0.608 | 0.588 | 0.688 | 0.674 | 0.659 | 0.655 |
| R-CBC (DeepCBC) | 0.664 | 0.624 | 0.592 | 0.675 | 0.677 | 0.679 | 0.659 |

**Table A4:** Model performance metrics by trial arm in non-anaemic population ([Sex][Donation interval in weeks]). ROC AUC, receiver operator characteristic area under the curve; AP, average precision; PPV, positive predictive value.

| Model | Trial Arm |  |  |  |  |  |  |
| --- | --- | --- | --- | --- | --- | --- | --- |
|  | M08 | M10 | M12 | F12 | F14 | F16 | All |
| **ROC AUC** |  |  |  |  |  |  |  |
| EHR-CBC (XGBoost) | 0.875 | 0.857 | 0.859 | 0.831 | 0.811 | 0.831 | 0.850 |
| HD-CBC (XGBoost) | 0.911 | 0.905 | 0.899 | 0.881 | 0.869 | 0.879 | 0.895 |
| R-CBC (DeepCBC) | 0.924 | 0.919 | 0.895 | 0.872 | 0.872 | 0.882 | 0.896 |
| **AP** |  |  |  |  |  |  |  |
| EHR-CBC (XGBoost) | 0.561 | 0.496 | 0.458 | 0.582 | 0.585 | 0.572 | 0.553 |
| HD-CBC (XGBoost) | 0.633 | 0.635 | 0.532 | 0.691 | 0.694 | 0.681 | 0.660 |
| R-CBC (DeepCBC) | 0.676 | 0.663 | 0.507 | 0.695 | 0.697 | 0.691 | 0.669 |
| **Balanced Accuracy** |  |  |  |  |  |  |  |
| EHR-CBC (XGBoost) | 0.786 | 0.782 | 0.801 | 0.750 | 0.742 | 0.749 | 0.774 |
| HD-CBC (XGBoost) | 0.831 | 0.812 | 0.812 | 0.809 | 0.785 | 0.795 | 0.811 |
| R-CBC (DeepCBC) | 0.834 | 0.831 | 0.810 | 0.793 | 0.791 | 0.817 | 0.817 |
| **Sensitivity** |  |  |  |  |  |  |  |
| EHR-CBC (XGBoost) | 0.730 | 0.716 | 0.738 | 0.770 | 0.791 | 0.785 | 0.762 |
| HD-CBC (XGBoost) | 0.793 | 0.750 | 0.723 | 0.803 | 0.791 | 0.765 | 0.776 |
| R-CBC (DeepCBC) | 0.805 | 0.796 | 0.723 | 0.784 | 0.818 | 0.806 | 0.793 |
| **Specificity** |  |  |  |  |  |  |  |
| EHR-CBC (XGBoost) | 0.842 | 0.847 | 0.864 | 0.730 | 0.693 | 0.712 | 0.787 |
| HD-CBC (XGBoost) | 0.869 | 0.875 | 0.901 | 0.816 | 0.778 | 0.824 | 0.847 |
| R-CBC (DeepCBC) | 0.863 | 0.867 | 0.897 | 0.801 | 0.764 | 0.828 | 0.841 |
| **PPV** |  |  |  |  |  |  |  |
| EHR-CBC (XGBoost) | 0.436 | 0.394 | 0.360 | 0.442 | 0.428 | 0.394 | 0.412 |
| HD-CBC (XGBoost) | 0.504 | 0.453 | 0.429 | 0.548 | 0.509 | 0.509 | 0.499 |
| R-CBC (DeepCBC) | 0.496 | 0.452 | 0.423 | 0.525 | 0.502 | 0.528 | 0.495 |
| **F1** |  |  |  |  |  |  |  |
| EHR-CBC (XGBoost) | 0.546 | 0.508 | 0.484 | 0.562 | 0.556 | 0.524 | 0.535 |
| HD-CBC (XGBoost) | 0.616 | 0.565 | 0.539 | 0.652 | 0.619 | 0.612 | 0.607 |
| R-CBC (DeepCBC) | 0.614 | 0.576 | 0.534 | 0.629 | 0.622 | 0.638 | 0.609 |

### A.4 DeepCBC predictions: Investigation of misclassified samples

We evaluated DeepCBC’s prediction accuracy over parameter ranges in haemoglobin, MCV, MCH, and ferritin (Fig. A4a). A decrease to near random chance (0.54) could be observed within a ferritin range of 15–20 µg*/*L compared to samples with ferritin *<* 10 µg*/*L and *>* 39 µg*/*L which were classified with over 90 % accuracy. Between 20– 39 µg*/*L, classification accuracy rose steadily.

Focusing further on model performance by ferritin range, we observed that the model output probability for ID rises with decreasing ferritin. This rise is especially steep for a ferritin level *<* 30 µg*/*L (Fig. A4b).

**Fig. A4:**
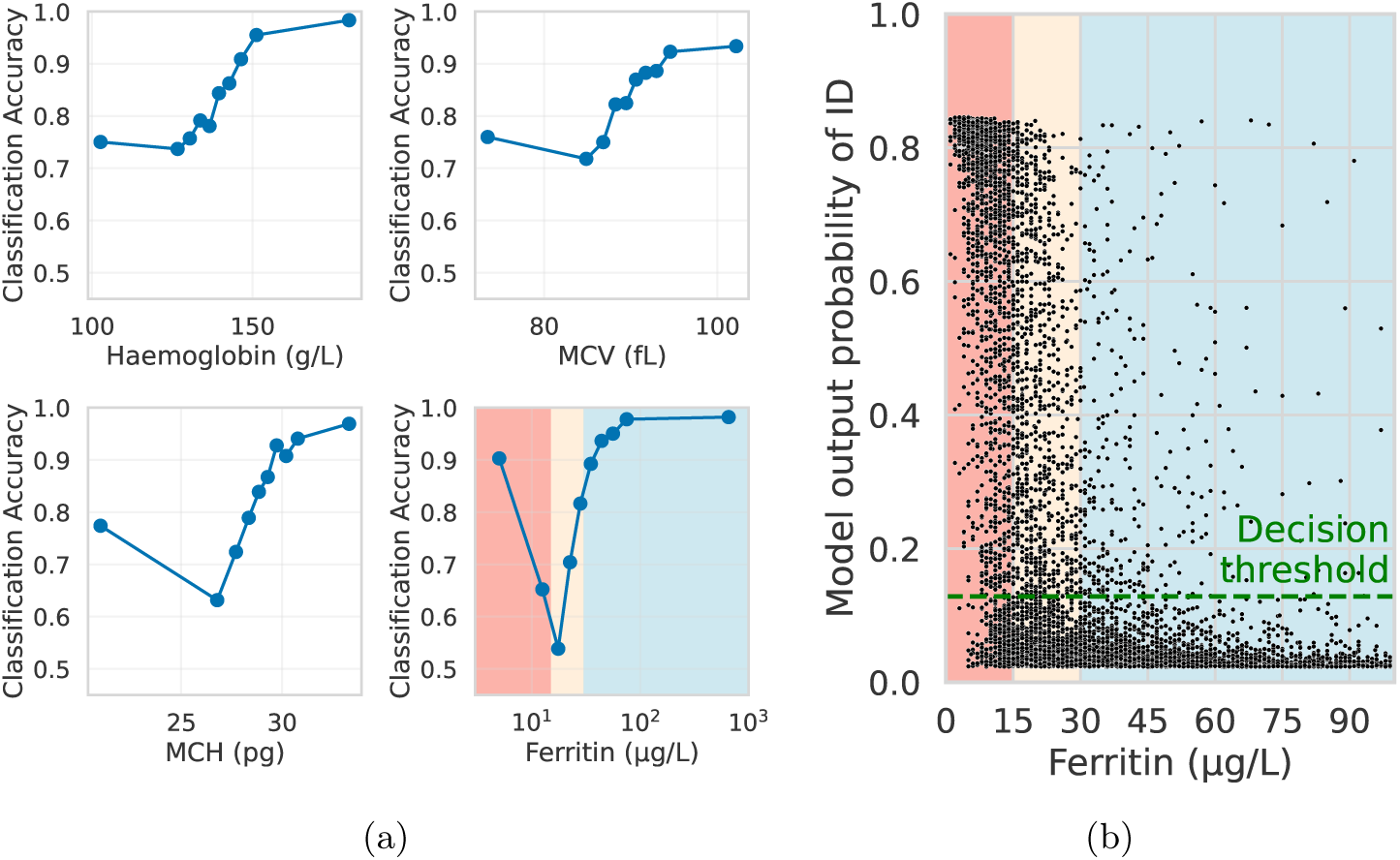
DeepCBC prediction behaviour over haemoglobin, MCV, MCH, and ferritin. **a,** Model classification accuracy across haemoglobin, MCV, MCH, and ferritin ranges. Background colours indicate ID (red), low iron stores (orange), and iron replete (blue) ferritin ranges. Points positioned at deciles of the corresponding parameter distribution. **b,** Model output probabilities for ID plotted against ferritin values in the test set, with background colouring as in (**a**). The green dashed line indicates the model’s decision threshold.

### A.5 DeepCBC predictions: haemoglobin over iron measurements

Extended Data Fig. A5b demonstrates that positive model predictions align with alterations in multiple iron markers beyond ferritin alone. The model’s positive predictions corresponded to samples with median transferrin saturation of 27.6 % vs 40.5 % for negative predictions, median hepcidin of 2.1 ng*/*mL vs 9.3 ng*/*mL, median soluble transferrin receptor (sTfR) of 3.9 mg*/*L vs 2.9 mg*/*L, and median sTfR */* log(ferritin) ratio of 3.3 vs 1.7 (all p *<* 0.0001, Mann–Whitney *U* test).

**Fig. A5:**
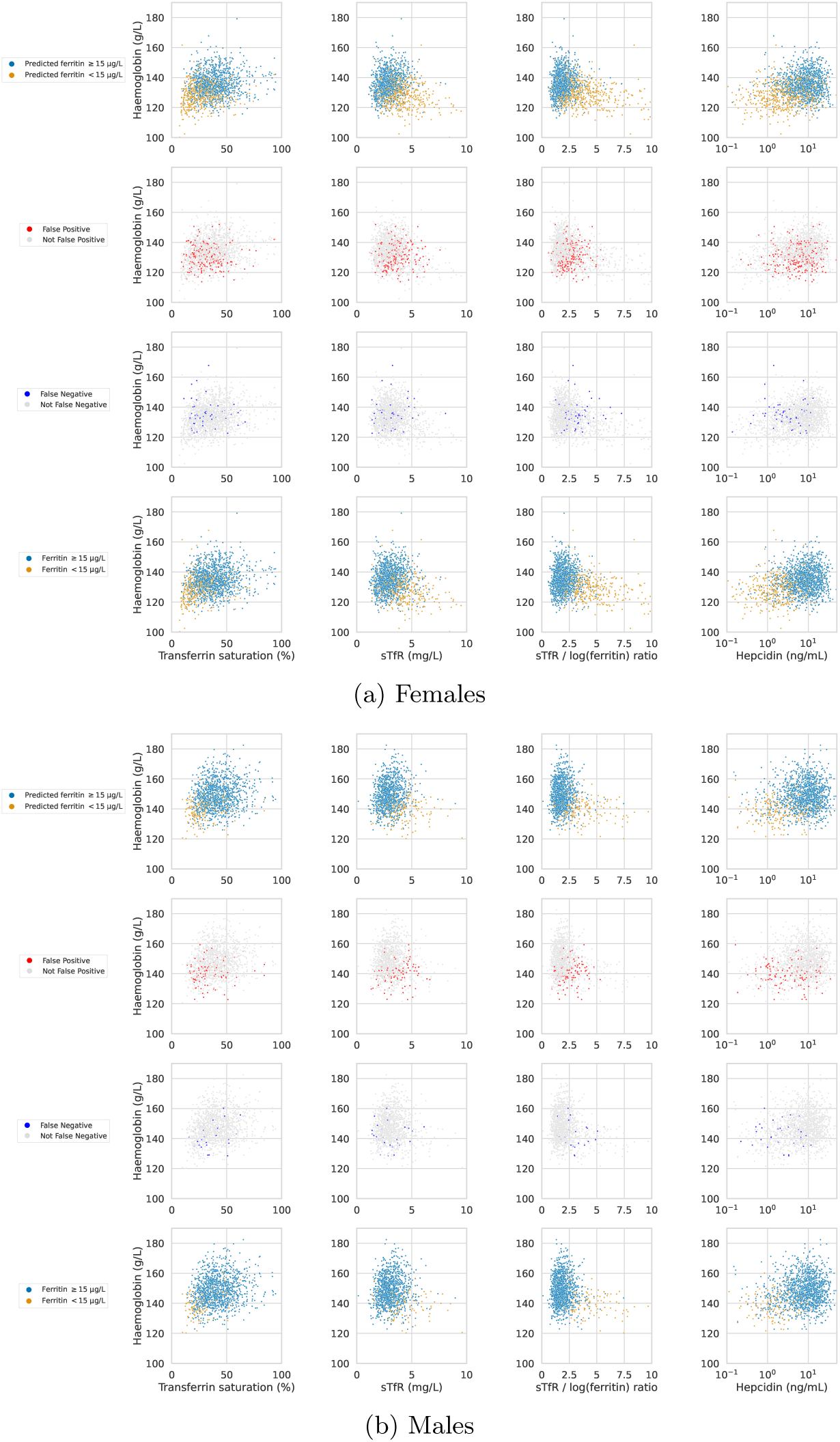
Scatter plots of haemoglobin over transferrin saturation, hepcidin concentration, sTfR, and sTfR to log ferritin ratio measurements for subjects in the test set (top) and INTERVAL baseline (bottom), coloured by DeepCBC model prediction and measured ferritin range, respectively.

### A.6 Ablation of RET and PLT-F channel and HD-CBC components for ID detection

We sought to quantify the incremental gain with increasing data granularity. Models tested were XGBoost models using EHR-CBC data, HD-CBC data without R-CBC-derived Gaussian parameters and IP messages and quality flags, HD-CBC with only the Gaussian parameters removed, HD-CBC, and DeepCBC using R-CBC data. We performed four separate experiments: using all channels in CBC measurements, excluding PLT-F channel, excluding RET channel, and excluding both PLT-F and RET channels (Extended Data Fig. A6). We observed progressive performance gains from the EHR-CBC model (conventional CBC features alone), through iterations of the HD-CBC model comprising extended summary reports, then with additions of IP messages and quality flags, to the full HD-CBC model including engineered Gaussian distribution features extracted from cell populations in the R-CBC. The best performance was achieved using R-CBC data input into the DeepCBC model. Channel ablations demonstrate that the RET channel particularly enhances performance for ID detection.

**Fig. A6:**
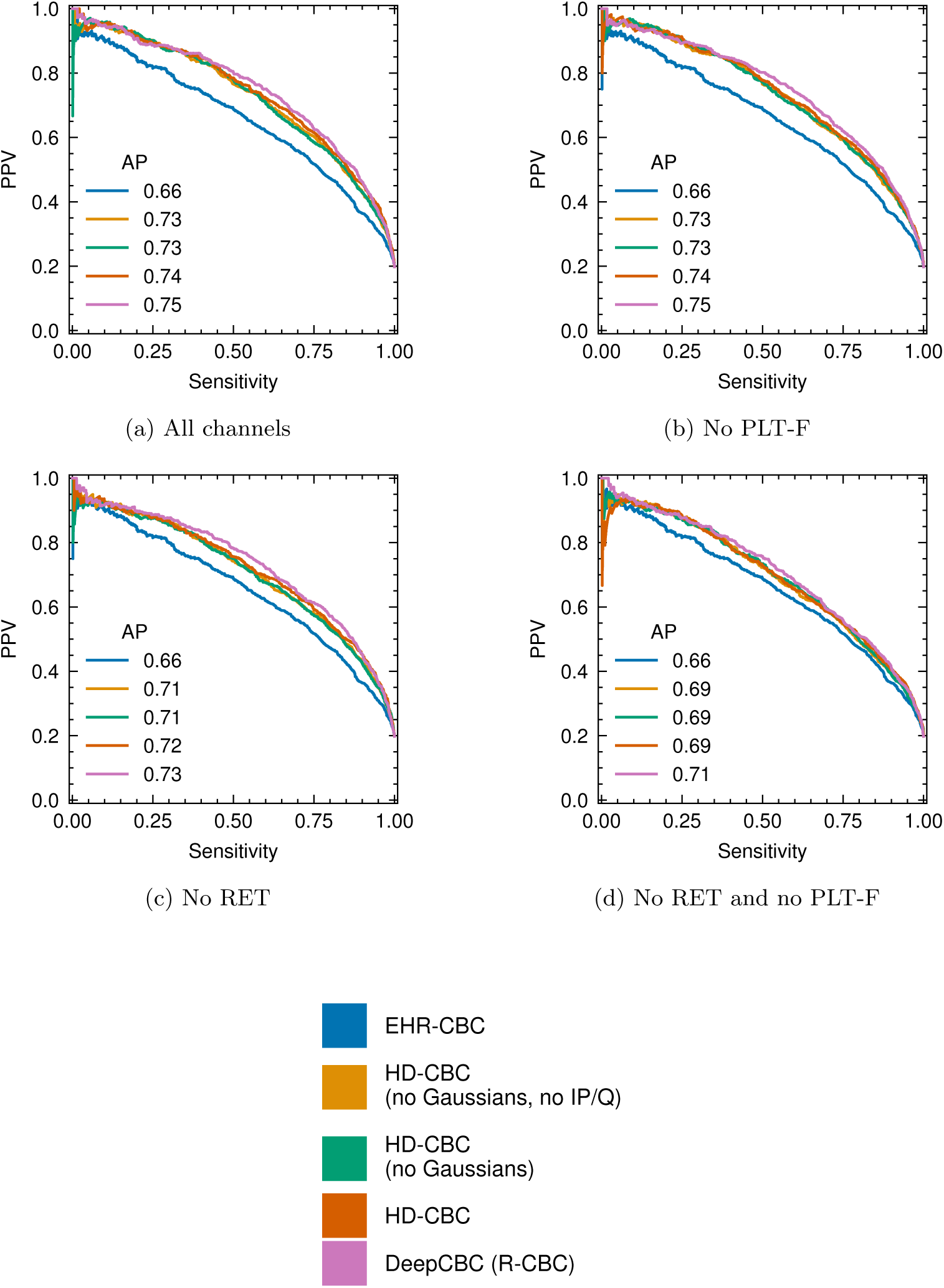
Channel and feature ablation for ID detection task. PPV-Sensitivity curves for detection of ferritin *<* 15 µg*/*L in the INTERVAL test set.

